# Knowledge barriers in the symptomatic-COVID-19 testing programme in the UK: an observational study

**DOI:** 10.1101/2021.03.16.21253719

**Authors:** Mark S. Graham, Anna May, Thomas Varsavsky, Carole H. Sudre, Benjamin Murray, Kerstin Kläser, Michela Antonelli, Liane S. Canas, Erika Molteni, Marc Modat, M. Jorge Cardoso, David A. Drew, Long H. Nguyen, Benjamin Rader, Christina Hu, Joan Capdevila, Alexander Hammers, Andrew T. Chan, Jonathan Wolf, John S. Brownstein, Tim D. Spector, Sebastien Ourselin, Claire J. Steves, Christina M. Astley

## Abstract

**Background:** Symptomatic testing programmes are crucial to the COVID-19 pandemic response. We sought to examine United Kingdom (UK) testing rates amongst individuals with test-qualifying symptoms, and factors associated with not testing.

**Methods:** We analysed a cohort of untested symptomatic app users (N=1,237), nested in the Zoe COVID Symptom Study (Zoe, N= 4,394,948); and symptomatic survey respondents who wanted, but did not have a test (N=1,956), drawn from the University of Maryland-Facebook Covid-19 Symptom Survey (UMD-Facebook, N=775,746).

**Findings:** The proportion tested among individuals with incident test-qualifying symptoms rose from ∼20% to ∼75% from April to December 2020 in Zoe. Testing was lower with one vs more symptoms (73.0% vs 85.0%), or short vs long symptom duration (72.6% vs 87.8%). 40.4% of survey respondents did not identify all three test-qualifying symptoms. Symptom identification decreased for every decade older (OR=0.908 [95% CI 0.883-0.933]). Amongst symptomatic UMD-Facebook respondents who wanted but did not have a test, not knowing where to go was the most cited factor (32.4%); this increased for each decade older (OR=1.207 [1.129-1.292]) and for every 4-years fewer in education (OR=0.685 [0.599-0.783]).

**Interpretation:** Despite current UK messaging on COVID-19 testing, there is a knowledge gap about when and where to test, and this may be contributing to the ∼25% testing gap. Risk factors, including older age and less education, highlight potential opportunities to tailor public health messages.

**Funding:** Zoe Global Limited, Department of Health, Wellcome Trust, EPSRC, NIHR, MRC, Alzheimer’s Society, Facebook Sponsored Research Agreement.

**Research in context:** *Evidence before this study:* To assess current evidence on test uptake in symptomatic testing programmes, and the reasons for not testing, we searched PubMed from database inception for research using the keywords (COVID-19) AND (testing) AND ((access) OR (uptake)). We did not find any work reporting on levels of test uptake amongst symptomatic individuals. We found three papers investigating geographic barriers to testing. We found one US based survey reporting on knowledge barriers to testing, and one UK based survey reporting on barriers in the period March - August 2020. Neither of these studies were able to combine testing behaviour with prospectively collected symptom reports from the users surveyed.

*Added value of this study:* Through prospective collection of symptom and test reports, we were able to estimate testing uptake amongst individuals with test-qualifying symptoms in the UK. Our results indicate that whilst testing has improved since the start of the pandemic, there remains a considerable testing gap. Investigating this gap we find that individuals with just one test-qualifying symptom or short symptom duration are less likely to get tested. We also find knowledge barriers to testing: a substantial proportion of individuals do not know which symptoms qualify them for a COVID-19 test, and do not know where to seek testing. We find a larger knowledge gap in individuals with older age and fewer years of education.

*Implications of all the available evidence:* Despite the UK having a simple set of symptom-based testing criteria, with tests made freely available through nationalised healthcare, a quarter of individuals with qualifying symptoms do not get tested. Our findings suggest testing uptake may be limited by individuals not acting on mild or transient symptoms, not recognising the testing criteria, and not knowing where to get tested. Improved messaging may help address this testing gap, with opportunities to target individuals of older age or fewer years of education. Messaging may prove even more valuable in countries with more fragmented testing infrastructure or more nuanced testing criteria, where knowledge barriers are likely to be greater.

## Introduction

Testing is a crucial component of the COVID-19 public health response to guide mitigation and triage illness, even as countries roll out vaccination campaigns. Whilst mass, population-based testing has been trialled,^1–3^ the majority of programmes seek to test individuals experiencing a certain set of symptoms. A successful program needs high testing uptake among those with test-qualifying symptoms.^4^ Achieving high uptake requires both an informed and willing population, and sufficient infrastructure to ensure test availability and accessibility.

In the United Kingdom (UK), the test-qualifying symptoms (fever, cough, or loss of smell)^5^ are relatively straightforward, have been consistent since loss of smell was added to the criteria on 18 May 2020, and are buttressed by a free, high-capacity, national testing programme. This is in contrast to other countries where criteria for testing have been more nuanced, varied over time and between regions, and testing access remains suboptimal.^6, 7^ Yet, despite the strengths of the UK programme, we observed that 25% of symptom-tracking app participants do not report testing despite experiencing test-qualifying symptoms. This and other evidence^8, 9^ raised questions regarding how the path from symptoms to testing could be improved to fully support the pandemic response.

Prior research focuses on logistical barriers for not getting tested such as geographic, socioeconomic and structural disparities in testing access,^6, 8, 10^ but there are other important barriers, including the knowledge required to successfully navigate the journey from symptom onset to test completion. Examining the reasons why people do not complete testing is hindered by the difficulty in identifying individuals who should have, but did not, receive COVID-19 tests.

Towards this end, we leveraged longitudinal data from over 4 million Zoe COVID Symptom Study (Zoe) participants,^11^ and over 700,000 surveys from the University of Maryland-Facebook Covid-19 Symptom Survey (UMD-Facebook), to describe the temporal changes in COVID-19 testing among UK residents with test-qualifying symptoms. We followed-up with cross-sectional surveys of the untested to identify knowledge barriers along the full journey to successful testing.

## Methods

This research combines syndromic surveillance data from the UK Zoe COVID Symptom Study (Zoe)^11^ and the UK UMD-Facebook COVID-19 Symptom Survey (UMD-Facebook)^12^ Additionally, more detailed follow-up surveys of recently untested symptomatic participants were analyzed. Survey details provided in Supplement S1-S3. Throughout we define test-qualifying symptoms using the UK’s National Health Service (NHS) criteria: high temperature; new, continuous cough; or loss or change to sense of smell or taste.^5^

### Data sources

#### UK Zoe COVID Symptom Study (Zoe)

Longitudinal data were prospectively collected using the Zoe COVID Symptom Study app, developed by Zoe Global with input from King’s College London (UK), the Massachusetts General Hospital (Boston, USA), and Lund and Uppsala Universities (Sweden). We used data from app launch on 24 March 2020 through 1 January 2021 (N=4,394,948, n=245,505,763 user-reports). App details are published elsewhere.^11^ Briefly, participants are asked enrollment questions at baseline, and then daily whether they feel physically normal or if they are experiencing symptoms. Participants are asked to record all COVID-19 test dates, types, and outcomes. To support COVID-19 incidence estimation,^13^ from 28 April 2020 the UK Department of Health and Social Care (DHSC) allocated polymerase chain reaction (PCR) tests to participants reporting any symptom after ≥1 “well” report in 9 days.

#### UK Zoe Follow-up Survey

To better understand the reasons why individuals who experience test-qualifying symptoms do not get tested, we deployed a cross-sectional SurveyMonkey web survey. We targeted participants reporting ≥1 test-qualifying symptoms for the first time between 14 November and 8 December 2020, who did not have a COVID-19 swab test report −7 to +14 days from symptom onset, including data entered up through 15 December 2020. Survey responses were linked to Zoe accounts using a unique, anonymised, user identifier. There were four survey sections to assess test-qualifying symptom recall and recognition, and test seeking and access. The survey was refined based on analysis of N=194 pilot survey responses sent to N=1,000. On 18 December 2020, the final survey was delivered by email to eligible participants (N=4,936 less N=706 without valid email address).

#### UK University of Maryland-Facebook COVID-19 Symptom Survey (UMD-Facebook)

This research is based on survey results from the University of Maryland (UMD).^12^ UMD, in collaboration with Facebook, delivered web-based, cross-sectional surveys to users sampled from the Facebook active user base. Survey sampling strategies were used to increase representativeness of the source population (here, the UK population) by sampling from the UK Facebook active user base and raking across census age, sex and geographic region to develop survey weights.^14^ The study was drawn from N=775,746 responses within UK geographic regions from 30 April 30 2020 (launch) through 21 February 2021. Primary analyses use raw data, and sensitivity analysis applied survey weights.

#### UMD-Facebook Symptomatic Never Tested but Desired Testing Survey Subcohort

On December 21, 2020, additional survey questions were asked of the “never tested” UMD-Facebook respondents regarding whether they had wanted to test in the prior two weeks, and reasons for not getting a test when they wanted one. For the analysis of knowledge-based factors contributing to not getting a test, cross-sectional surveys were limited to a subcohort of those surveys completed from December 21, 2020 onwards (N=205,017, survey versions 7-9), reporting test-qualifying symptoms in the past 24 hours (N=32,711, 16.0%), reported having never been tested for coronavirus (N=12,821, 39.2%) and reported having wanted testing in the prior 14 days (N=1,956, 15.3%). To describe the factors associated with knowledge barriers to successful testing, we focused on the question “Do any of the following reasons describe why you haven’t been tested for coronavirus (COVID-19) in the last X days?”, where X is symptom duration up to 14 days, and the response option “I don’t know where to go”.

#### Data analysis

We calculated the proportions of outcomes among subgroups, considering several outcomes and subgroup definitions. Logistic regression was used to estimate the covariate-outcome association. Covariates considered varied for each analysis as not all variables available in each data set: sex, age, symptom (see Supplementary Table 4), symptom number and duration, symptom-to-survey time, self-reported years of education, index of multiple deprivation [IMD]^15^, profession/work, self-reported or national rural-urban classification [RUC])^16^. For some analyses, Zoe reports of either loss of taste/smell or altered taste/smell were combined.^16^ Zoe and UMD-Facebook analyses were conducted using Python 3.8 and R 3.6.3, respectively.

## Results

### Quantifying the Testing Gap

The proportion of Zoe participants reporting COVID-19 testing has increased over time (Figure 1), from <20% (1 April 2020) to >70% (1 January 2021). In mid-September, national PCR testing capacity was exceeded,^17^ coincident with a transient drop in reported tests. In late 2020, despite adequate test capacity, >25% of test-qualifying app users did not report a test. The UMD-Facebook survey time-series mirrors these trends (Supplementary Figure 5), albeit with a lower absolute proportion, likely due, in part, to survey design differences (e.g. ever tests and symptoms in prior 24 hours, vs tests −7 to +14 days from incident symptoms). The UMD-Facebook testing gap is generally lower amongst respondents with a smartphone, and even lower among symptom-tracking app participants (not necessarily Zoe).

**Figure 1.**
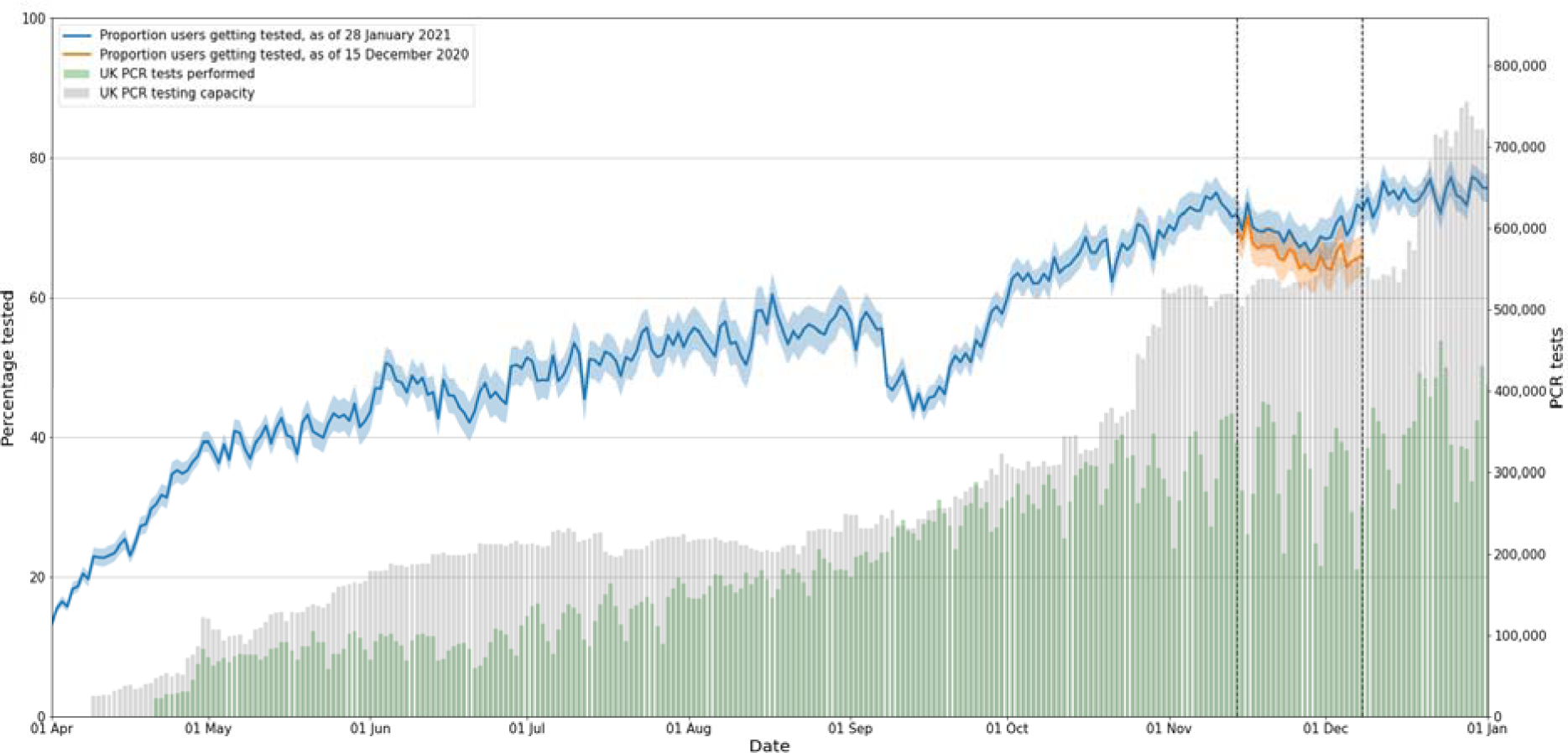
Proportion of participants reporting at least one test-qualifying symptom for the first time that logged a COVID-19 swab test −7 to +14 days after the onset of the symptom. Dashed vertical lines indicate the three-week study period (14 November 2020 to 8 December 2020) that was used to define participants eligible for a follow-up survey, i.e. those that reported test-qualifying symptoms for the first time but no swab test were invited to the Zoe Follow-Up Survey. Blue line calculated using data logged by 28 January 2020, red line calculated using data logged by 15 December 2020, which was the dataset used to identify target participants for the survey. The difference shows that some participants recorded test data after selection into the follow-up survey cohort. Grey bars indicate the UK PCR testing capacity, while green bars indicate PCR tests performed.

### Symptom Severity and Not Testing

To better understand the factors contributing to COVID-19 testing, Zoe participants with test-qualifying symptoms during the study period, who did not report testing (N=20,425), were studied further. During this period, the proportion not tested among those with test-qualifying symptoms was higher for those with 1 vs ≥2 test-qualifying symptoms (27.0% vs 15.0%), and for those with symptoms lasting ≤2 days vs >2 (27.4% vs 12.2%), (Table 1). Similarly, the proportion of ever-tested in UMD-Facebook was lowest among those with only one test-qualifying symptom or short symptom duration (Supplementary Figure 5). A total of 1,254 users (26.6%) responded to the follow-up survey. Zoe and follow-up survey participants during this period were younger and more female than the general population, similar to the demographic trends reported previously in Zoe^13^ and other digital health studies (Supplement S6).^18, 19^

**Table 1.**
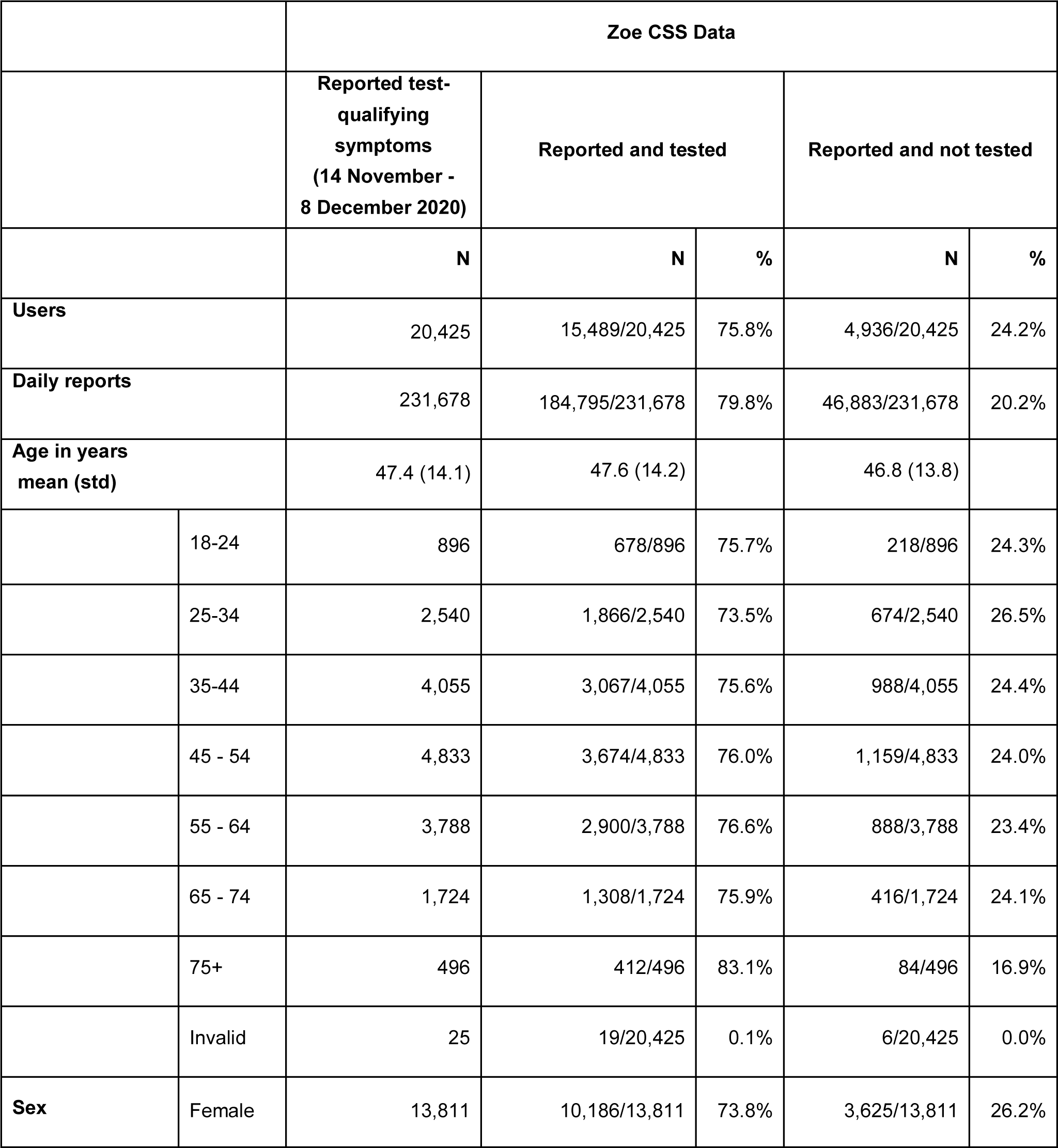
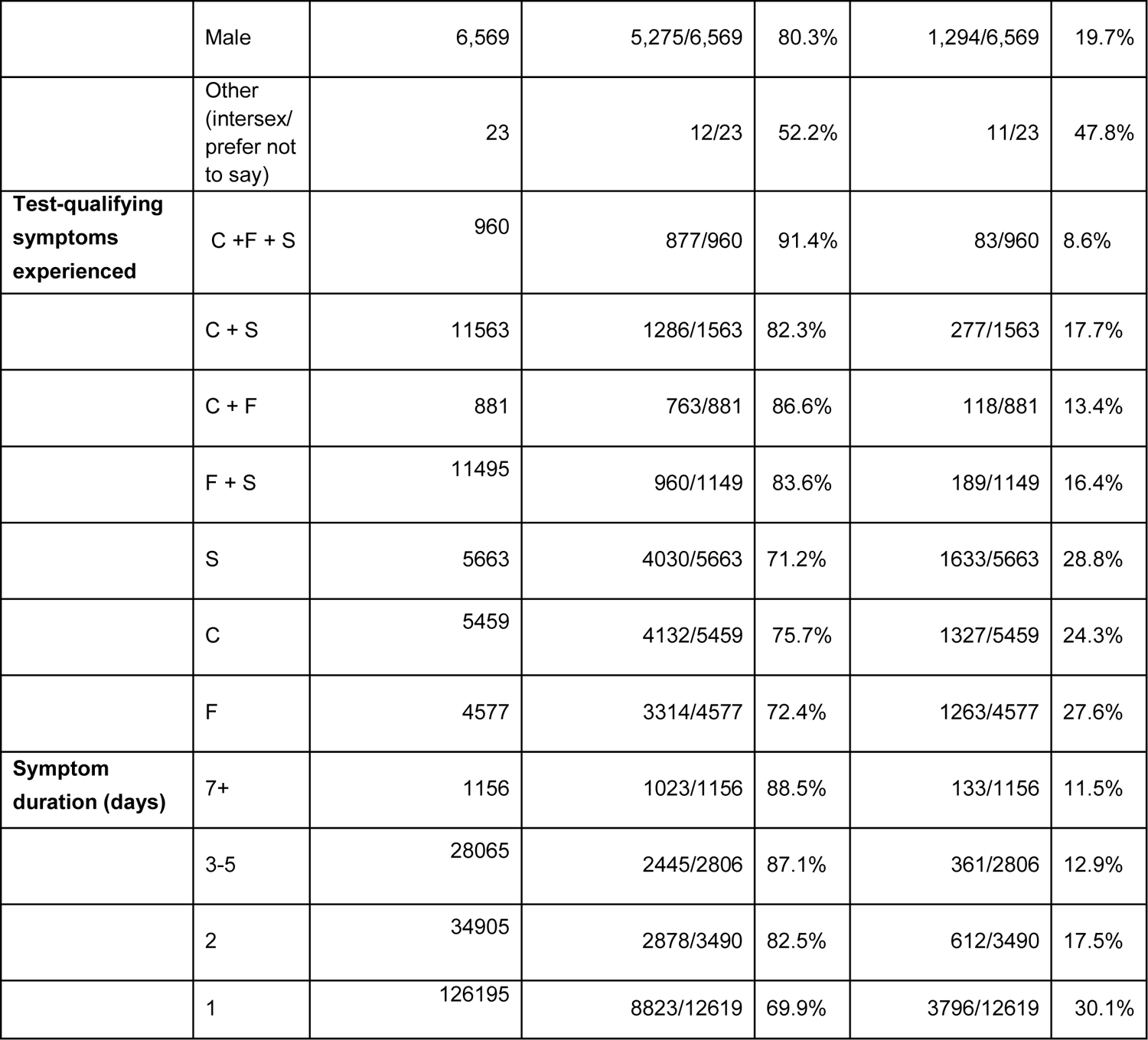
Characteristics of users reporting test-qualifying symptoms for the first time between 14 November and 8 December 2020, inclusive for Zoe CSS App users. Symptom key: F=fever, C=persistent cough, S=loss or altered sense of taste or smell.

|  |  | Zoe CSS Data |  |  |  |  |
| --- | --- | --- | --- | --- | --- | --- |
|  |  | Reported test-qualifying symptoms<br>(14 November - 8 December 2020) | Reported and tested |  | Reported and not tested |  |
|  |  | N | N | % | N | % |
| **Users** |  | 20,425 | 15,489/20,425 | 75.8% | 4,936/20,425 | 24.2% |
| **Daily reports** |  | 231,678 | 184,795/231,678 | 79.8% | 46,883/231,678 | 20.2% |
| **Age in years mean (std)** |  | 47.4 (14.1) | 47.6 (14.2) |  | 46.8 (13.8) |  |
|  | 18-24 | 896 | 678/896 | 75.7% | 218/896 | 24.3% |
|  | 25-34 | 2,540 | 1,866/2,540 | 73.5% | 674/2,540 | 26.5% |
|  | 35-44 | 4,055 | 3,067/4,055 | 75.6% | 988/4,055 | 24.4% |
|  | 45 - 54 | 4,833 | 3,674/4,833 | 76.0% | 1,159/4,833 | 24.0% |
|  | 55 - 64 | 3,788 | 2,900/3,788 | 76.6% | 888/3,788 | 23.4% |
|  | 65 - 74 | 1,724 | 1,308/1,724 | 75.9% | 416/1,724 | 24.1% |
|  | 75+ | 496 | 412/496 | 83.1% | 84/496 | 16.9% |
|  | Invalid | 25 | 19/20,425 | 0.1% | 6/20,425 | 0.0% |
| **Sex** | Female | 13,811 | 10,186/13,811 | 73.8% | 3,625/13,811 | 26.2% |
|  | Male | 6,569 | 5,275/6,569 | 80.3% | 1,294/6,569 | 19.7% |
|  | Other<br>(intersex/<br>prefer not<br>to say) | 23 | 12/23 | 52.2% | 11/23 | 47.8% |
| **Test-qualifying**<br>**symptoms**<br>**experienced** | C + F + S | 960 | 877/960 | 91.4% | 83/960 | 8.6% |
|  | C + S | 11563 | 1286/1563 | 82.3% | 277/1563 | 17.7% |
|  | C + F | 881 | 763/881 | 86.6% | 118/881 | 13.4% |
|  | F + S | 11495 | 960/1149 | 83.6% | 189/1149 | 16.4% |
|  | S | 5663 | 4030/5663 | 71.2% | 1633/5663 | 28.8% |
|  | C | 5459 | 4132/5459 | 75.7% | 1327/5459 | 24.3% |
|  | F | 4577 | 3314/4577 | 72.4% | 1263/4577 | 27.6% |
| **Symptom**<br>**duration (days)** | 7+ | 1156 | 1023/1156 | 88.5% | 133/1156 | 11.5% |
|  | 3-5 | 28065 | 2445/2806 | 87.1% | 361/2806 | 12.9% |
|  | 2 | 34905 | 2878/3490 | 82.5% | 612/3490 | 17.5% |
|  | 1 | 126195 | 8823/12619 | 69.9% | 3796/12619 | 30.1% |

### Journey to Successful COVID-19 Testing

In the Zoe follow-up survey, only 42.1% survey respondents recalled having experienced at least one test-qualifying symptom in the past month (Table 2). Of those who recalled their symptoms, 54.7% recognised that these symptoms qualified them for a COVID-19 test. Among participants who recognized test-qualifying symptoms, they were likely to go on to attempt (85.6%) and then successfully obtain a COVID-19 test (93.0%, or 18.4% of all survey respondents).

**Table 2.** Overall survey responses by stage of the testing journey from recall of symptoms logged, recognition of symptoms requiring testing, to attempting to test and lastly reporting a test.

|  | **Stage of testing journey** |  |  |  |
| --- | --- | --- | --- | --- |
|  | **Recalled**<br>**symptoms** | **Recognised**<br>**symptoms**<br>**required testing** | **Attempted**<br>**testing** | **Succeeded** |
| **All** | 521/1237<br>(42.1%) | 285/521<br>(54.7%) | 244/285<br>(85.6%) | 227/244<br>(93.0%) |

### Recall of Previously-Reported Test-Qualifying Symptoms

Among those who reported a single test-qualifying symptom (Table 3), recall of absent or altered taste/smell (45.8%) and cough (40.8%), was higher than fever (25.5%), the last of which, as queried in-app (“Do you have a fever or feel too hot?”) was an admixture of two symptoms that may not be recalled equally well. Recall of having experienced test-qualifying symptoms was associated with number of symptoms experienced (per test-qualifying symptom OR=1.302 [95% CI 1.220 - 1.391), symptom duration (per day OR=1.065 [95% CI 1.054 - 1.076]) and recency (per symptom-to-survey days OR=0.995 [95% CI 0.991 - 1.000]). Number of symptoms and symptom duration remained significantly associated with recall after adjusting for age, sex, and recency of symptom onset (model results in Supplement S7).

**Table 3.** Recall of test-qualifying symptoms stratified by type of symptom, symptom duration, and time passed between experiencing symptoms and participation in the Zoe Follow-Up Survey. Participants were eligible for the survey if no test was logged one week before or two weeks after onset of symptoms. Symptom key: F=fever, C=persistent cough, S=loss or altered sense of taste or smell.

|  |  | **Recalled having experienced a test-qualifying symptom** |
| --- | --- | --- |
| **All users surveyed** |  | 529/1254 (42.2%) |
| **By symptoms experienced** | C + F + S | 22/27 (81.5%) |
|  | C + S | 56/66 (84.8%) |
|  | C + F | 12/20 (60.0%) |
|  | F + S | 19/36 (52.8%) |
|  | S | 187/408 (45.8%) |
|  | C | 138/338 (40.8%) |
|  | F | 83/325 (25.5%) |
| **By symptom duration** | 7+ days | 64/69 (92.8%) |
|  | 3-5 | 84/111 (75.7%) |
|  | 2 | 91/184 (49.5%) |
|  | 1 | 274/863 (31.7%) |
| **By time from symptoms to survey response** | 7-14 days | 91/176 (51.7%) |
|  | 14 - 21 days | 160/383 (41.8%) |
|  | 21 - 28 | 139/359 (38.7%) |
|  | 28 + | 131/319 (41.1%) |

### Recognizing COVID-19 Test-Qualifying Symptoms

Just 54.7% of those who recalled experiencing test-qualifying symptoms indicated their symptoms qualified them for a COVID-19 test. We queried the remaining respondents (N=809, i.e. those who did not recall or who did not indicate the symptoms they recalled qualified them for testing) which symptoms would qualify them for a test. These respondents were similarly able to recognize fever (63.4%), cough (67.6%), and loss of smell (65.4%), and much less so for altered smell (29.7%). Only 59.6% identified the triad of fever, cough, and loss of smell as qualifying for a test, with lower recognition amongst the oldest age groups (Table 4). In univariate analyses, each decade older reduced the odds of recognizing the triad (OR = 0.908 [95% CI 0.883 - 0.933]). This finding remained largely unchanged after adjustment for sex, IMD, and rural-urban living. We found similar associations with the outcome of identifying each individual symptoms (model results in Supplement S8). No associations were found for sex, age, IMD and rural-/urban living.

**Table 4.** Understanding of symptoms as factors qualifying for COVID-19 testing by age group, among the 809 survey respondents who did not recall having experienced a test-qualifying symptoms and were asked “What symptoms qualify you for a covid test where you live?”.

| Group |  |  | Recognises symptom as testing-qualifying |  |  |  |
| --- | --- | --- | --- | --- | --- | --- |
|  |  | All of: fever, persistent cough, and loss of smell | Fever | Persistent cough | Loss of smell | Altered smell |
| All |  | 482/809<br>(59.6%) | 513/809<br>(63.4%) | 548/809<br>(67.7%) | 529/809<br>(65.4%) | 240/809<br>(29.7%) |
| Age | 18 - 24 | 3/4<br>(75.0%) | 3/4 (75.0%) | 4/4<br>(100.0%) | 4/4<br>(100.0%) | 3/4 (75.0%) |
|  | 25 - 34 | 39/49<br>(79.6%) | 40/49<br>(81.6%) | 41/49<br>(83.7%) | 40/49<br>(81.6%) | 21/49<br>(42.9%) |
|  | 35 - 44 | 99/135<br>(73.3%) | 102/135<br>(75.6%) | 113/135<br>(83.7%) | 109/135<br>(80.7%) | 38/135<br>(28.1%) |
|  | 45 - 54 | 143/217<br>(65.9%) | 152/217<br>(70.0%) | 160/217<br>(73.7%) | 156/217<br>(71.9%) | 66/217<br>(30.4%) |
|  | 55 - 64 | 118/223<br>(52.9%) | 129/223<br>(57.8%) | 137/223<br>(61.4%) | 132/223<br>(59.2%) | 67/223<br>(30.0%) |
|  | 65 - 74 | 72/150<br>(48.0%) | 79/150<br>(52.7%) | 82/150<br>(54.7%) | 78/150<br>(52.0%) | 39/150<br>(26.0%) |
|  | 75+ | 8/31<br>(25.8%) | 8/31<br>(25.8%) | 11/31<br>(35.5%) | 10/31<br>(32.3%) | 6/31<br>(19.4%) |

### Reasons for Not Testing Among Those Who Qualified For and Wanted a Test

In the follow-up survey, there were few respondents (N=17) who recognised their symptoms qualified them for testing, and attempted, but did not succeed at testing (Table 2). We therefore evaluated complementary data from a subcohort of UMD-Facebook respondents from 21 December 2020 to 21 February 2021, who endorsed test-qualifying symptoms, who had never tested, and who indicated “yes” to the question question “Have you wanted to get tested for coronavirus (COVID-19) at any time in the last 14 days?” (N=1,956, Table 5). Among those who wanted testing, “I don’t know where to go” was the most frequently selected option (32.4%). The other multi-choice reasons were: “I am unable to travel to a testing location” (29.1%), “I tried to get a test but was not able to get one” (25.6%), “I am worried about bad things happening to me or my family (including discrimination, government policies, and social stigma)” (18.4%), “I can’t afford the cost of the test” (17.9%), and “I don’t have time to get tested”(13.3%). Given the scope of this study, we have focused on the knowledge-based response, though we acknowledge the logistical barriers are important.

**Table 5.**
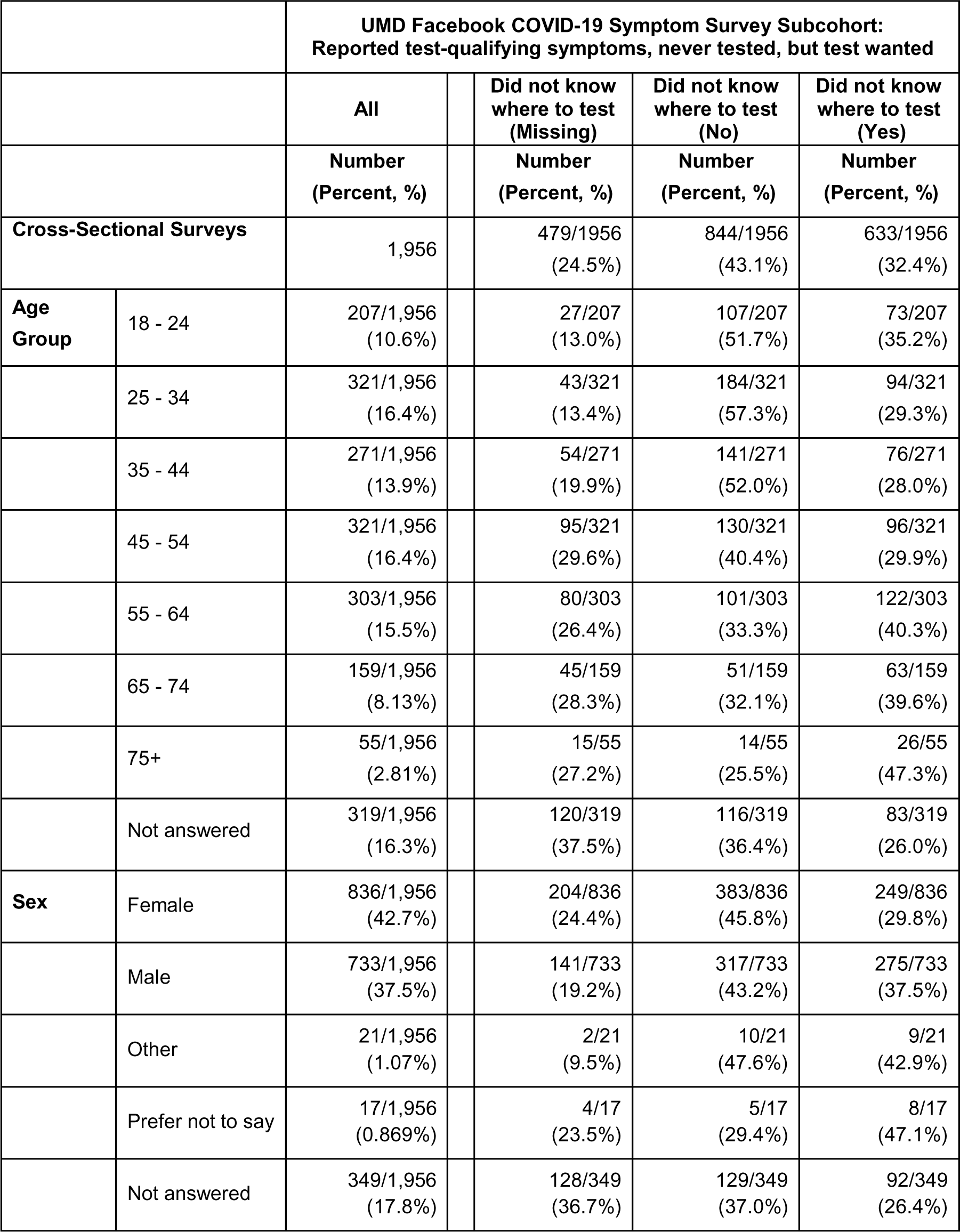
Characteristics of cross-sectional surveys between 21 December 2020 through February 21, 2021 for UMD-Facebook respondents reporting test-qualifying symptoms, never testing, but wanting to test in the prior 14 days. Survey-weighted mean age 39.3 years (unweighted 45.4 years) and proportion female of all male and female respondents 51.9%. Additional columns for the outcome of “I don’t know where to go” stratified by demographic factors, for those who responded “yes” “no”, and missing values.

|  |  | **UMD Facebook COVID-19 Symptom Survey Subcohort:**<br>**Reported test-qualifying symptoms, never tested, but test wanted** |  |  |  |
| --- | --- | --- | --- | --- | --- |
|  |  | **All** | **Did not know**<br>**where to test**<br>**(Missing)** | **Did not know**<br>**where to test**<br>**(No)** | **Did not know**<br>**where to test**<br>**(Yes)** |
|  |  | **Number**<br>**(Percent, %)** | **Number**<br>**(Percent, %)** | **Number**<br>**(Percent, %)** | **Number**<br>**(Percent, %)** |
| **Cross-Sectional Surveys** |  | 1,956 | 479/1956<br>(24.5%) | 844/1956<br>(43.1%) | 633/1956<br>(32.4%) |
| **Age**<br>**Group** | 18 - 24 | 207/1,956<br>(10.6%) | 27/207<br>(13.0%) | 107/207<br>(51.7%) | 73/207<br>(35.2%) |
|  | 25 - 34 | 321/1,956<br>(16.4%) | 43/321<br>(13.4%) | 184/321<br>(57.3%) | 94/321<br>(29.3%) |
|  | 35 - 44 | 271/1,956<br>(13.9%) | 54/271<br>(19.9%) | 141/271<br>(52.0%) | 76/271<br>(28.0%) |
|  | 45 - 54 | 321/1,956<br>(16.4%) | 95/321<br>(29.6%) | 130/321<br>(40.4%) | 96/321<br>(29.9%) |
|  | 55 - 64 | 303/1,956<br>(15.5%) | 80/303<br>(26.4%) | 101/303<br>(33.3%) | 122/303<br>(40.3%) |
|  | 65 - 74 | 159/1,956<br>(8.13%) | 45/159<br>(28.3%) | 51/159<br>(32.1%) | 63/159<br>(39.6%) |
|  | 75+ | 55/1,956<br>(2.81%) | 15/55<br>(27.2%) | 14/55<br>(25.5%) | 26/55<br>(47.3%) |
|  | Not answered | 319/1,956<br>(16.3%) | 120/319<br>(37.5%) | 116/319<br>(36.4%) | 83/319<br>(26.0%) |
| **Sex** | Female | 836/1,956<br>(42.7%) | 204/836<br>(24.4%) | 383/836<br>(45.8%) | 249/836<br>(29.8%) |
|  | Male | 733/1,956<br>(37.5%) | 141/733<br>(19.2%) | 317/733<br>(43.2%) | 275/733<br>(37.5%) |
|  | Other | 21/1,956<br>(1.07%) | 2/21<br>(9.5%) | 10/21<br>(47.6%) | 9/21<br>(42.9%) |
|  | Prefer not to say | 17/1,956<br>(0.869%) | 4/17<br>(23.5%) | 5/17<br>(29.4%) | 8/17<br>(47.1%) |
|  | Not answered | 349/1,956<br>(17.8%) | 128/349<br>(36.7%) | 129/349<br>(37.0%) | 92/349<br>(26.4%) |

### Not Knowing Where to Test Among the Symptomatic Wanting Testing

We further investigated demographic factors associated with not knowing where to go to obtain a test (Figure 2). Not knowing where to go to obtain a test (“yes” vs referent “no”) was associated with older age (per decade OR=1.207 [1.129-1.292]) and less education (per 4-years OR=0.685 [0.599-0.783]). Male sex (OR=1.334 [1.064-1.675]) and living outside a city (OR=1.201 [0.926-1.562]) were not significant with Bonferroni correction for multiple hypothesis testing (p-threshold 0.0125 = 0.05/4). Education was similarly protective for not knowing where to test adjusting for age and sex, use of survey weights, or assuming missing responses were “no” (models results in Supplementary S9).

**Figure 2.**
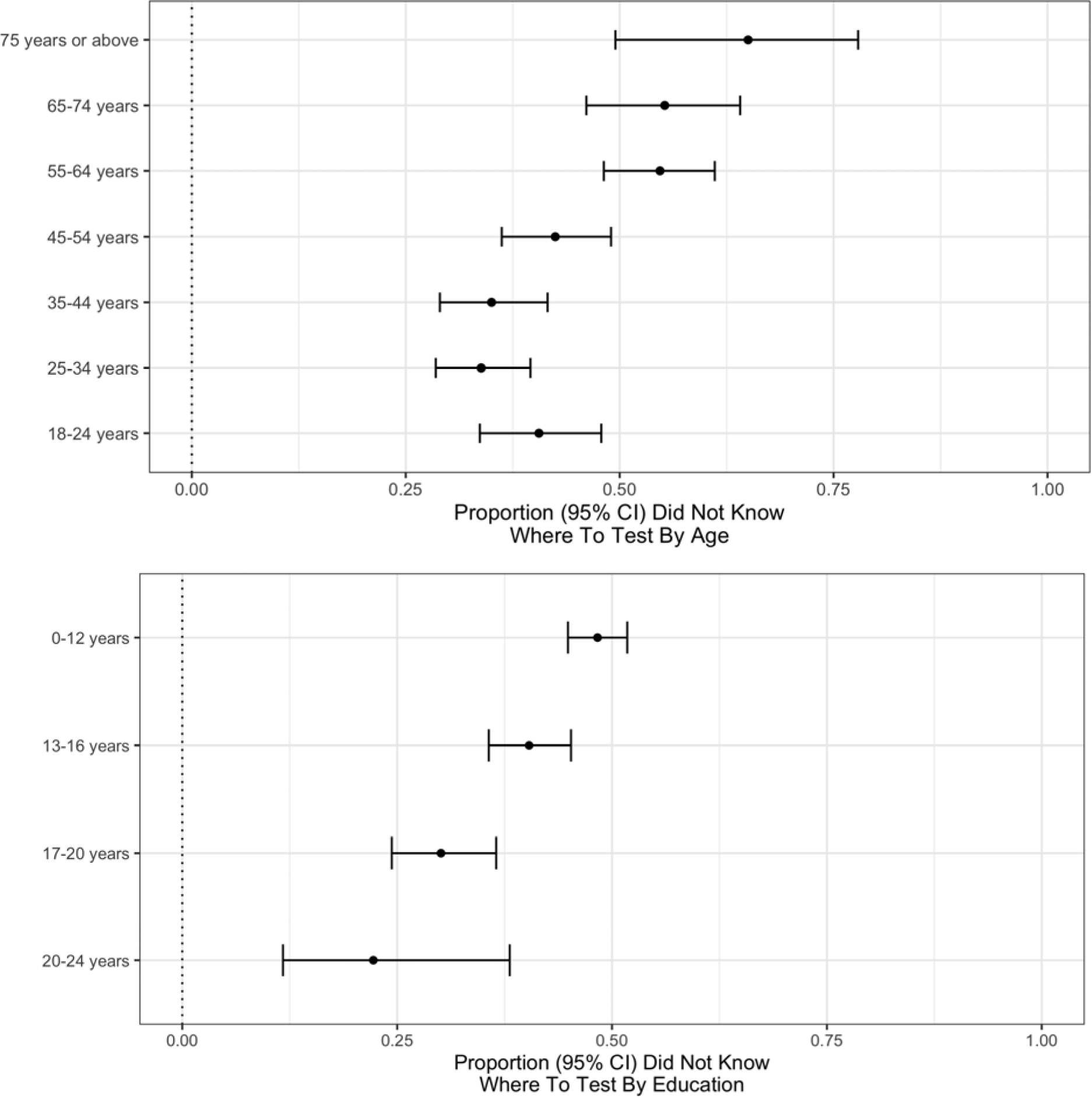
Proportion (95% confidence interval) of respondents who indicated “yes” (per “yes” plus “no”) for “I don’t know where to go” as a option to the question “Do any of the following reasons describe why you haven’t been tested for coronavirus (COVID-19) in the last X days?”, where X is the self-reported duration of symptom capped at 14 days. This was restricted to untested respondents with test-qualifying symptoms. Proportions stratified by age (top) and education (bottom).

Acknowledging our limited sample size, we conducted qualitative, hypothesis-generating analyses of other demographic factors correlated with knowledge barriers (Supplementary S10). While cities were not protective in the regression model, the proportion not knowing where to test was slightly lower in London than elsewhere (Supplementary S11). Having a smartphone and using a symptom-tracking app qualitatively had a bigger impact on the knowledge gap than the relatively small urban-rural and regional differences. There were modest qualitative differences in not knowing where to test by profession, with the highest proportions among those in transportation, tourism and construction and the lowest in finance, public administration and health.

## Discussion

### Persistent Testing Gap

Our analysis finds that in December 2020 approximately one quarter of symptomatic UK Zoe participants who qualified for a COVID-19 test did not undergo testing. The proportion of ever-tested recently-symptomatic UK UMD-Facebook respondents echoes this trend. While we show a substantial improvement from April, 2020, the persistent testing gap is problematic for pandemic management in the UK and elsewhere. Non-pharmaceutical mitigation strategies are likely to be required,^20–22^ despite effective vaccines, because of COVID-19 transmissibility and the anticipated time to reach herd immunity, even with a one-dose immunization strategy.^23^ The lower the proportion of identified infections, such as through insufficient testing of symptomatic cases, the more likely transmission events will go unchecked.

### Knowledge Barriers to Testing

With this testing gap in mind, we sought to characterize barriers to testing that might inform improvements to the UK testing programme using data from two large, complementary surveillance platforms. Through analysis of prospective self-reported testing outcomes, along with follow-up and population-sampled surveys, we identified three key knowledge barriers along the path to successful COVID-19 testing. Firstly, the association of less testing with brief and/or single test-qualifying symptoms suggests an implied severity threshold for testing that is inconsistent with guidance,^5^ and with the spectrum of infectious COVID-19.^24^ Individuals may minimise their symptoms or possibly hold the misconceived notion that COVID-19 manifests in a stereotypical manner.^25^ In a preprint DHSC report utilizing online surveys May 25 to August 5, 2020, those with test-qualifying symptoms did not request testing because symptoms were mild (16.0%), improved (16.1%), or they did not think symptoms were due to COVID-19 (20%).^8^

Secondly, we show that four of ten did not recognize all three of the triad of UK test-qualifying--i.e. fever, cough and loss of smell. The DHSC report^8^ estimated 51.1% of all respondents failed to recognize the triad. Despite increased media coverage and a second wave with mitigation intensification,^26^ recognition of these test-qualifying symptoms in our survey around six months later increased by ∼10% and therefore remains alarmingly low.

Lastly, one third of those who wanted a COVID-19 test cited not knowing where to test as a contributing factor to their not getting one. This was the most frequently cited reason among the six options. Thus, despite the fact that survey respondents are possibly more aware of health information, they acknowledge challenges to finding testing that are comparable to, if not greater than, logistical barriers to testing (e.g. travel, time), even within the framework of the more centralized UK testing programme.

### Public Health Implications

Our findings have significant public health implications. The UK NHS testing programme offers free COVID-19 tests to those with test-qualifying symptoms, with the list of qualifying symptoms unchanged since loss/alteration to taste and smell were included on 18 May 2020,^27^ and tests accessed through a central booking system.^5^ Risk mitigation and public health principles generally would agree with these key features of the UK program i.e. the use of concise and consistent guidance, and limiting logistical barriers to following guidance. In this sense, the UK is a sort of case study of the “best case scenario”, and yet there is still a significant gap in understanding. Not only are greater efforts needed to educate the UK public, it is likely that comparable efforts to mind the knowledge gap will be needed in countries with regionally varying testing criteria or methods of accessing testing.

Our work suggests there is a need for messaging improvements to the UK testing campaign. In our study, among the untested who qualified for a test, older age was associated with not knowing when and where to test. In the earlier DHSC report^8^, older age was generally protective with respect to testing knowledge and behaviors, perhaps suggesting knowledge gains in the young over the past six months. Fewer years of education was also associated with not knowing where to test in our study. We found suggestive evidence that the knowledge gap may be more pronounced among those who do not have smartphones. Older populations in pre-pandemic studies have slower adoption of certain technologies, yet the abrupt social isolation resulting from mitigation strategies may be leaving important segments of the population behind.^28^ Education attained and age are likely not the root cause. Rather they likely highlight pre-existing health-information disparities that have been exacerbated by a year of unprecedented changes in how individuals interface with each other and the world.

Our findings support the need for targeted messaging to certain at-risk demographic groups, possibly in a non-digital format (e.g. radio, community signage). Our findings are particularly timely in light of work showing that expansion of the symptoms that qualify for a test would help detect more cases, assuming those who qualify do indeed successfully test.^29, 30^ This theoretical gain in case detection could be lost if the change in tack leaves vulnerable populations behind. There is overlap between knowledge risk factors and COVID-19 risk, such as older age,^31^ though we did see a higher absolute rate of testing in the oldest age group. Overlap with vaccine hesitancy risk factors may further amplify disparities in healthcare access, leaving some groups both less tested and less protected.

Furthermore, messaging could also emphasise that even individuals with mild or transient symptoms may have COVID-19 and should get tested. COVID-19 has a broad spectrum of disease severity with a substantial number of cases being fully asymptomatic, and with asymptomatic carriers still being able to transmit, albeit at reduced rates.^24^

### Strengths and Limitations

The Zoe platform affords a unique opportunity to prospectively link testing behaviours with incident symptoms in a large user base comprising ∼6% of the UK population. The UMD-Facebook platform, though smaller in size and slightly different in survey design, corroborates temporal trends over in the broader population. To our knowledge, this has enabled the first time-varying estimate of testing rates amongst individuals that qualify for COVID-19 tests over the course of the pandemic. Both platforms could be leveraged to track the testing and knowledge gaps, in real time, allowing the effectiveness of interventions, such as improved messaging on when and where to test, to be assessed.

We acknowledge a number of limitations to this study. Digital surveys include selected populations not necessarily representative of the wider population. Such platforms have well-documented biases in demographic age, sex, and socioeconomic factors which we adjusted for in our analyses.^18, 32^ In addition, digital surveys may not be generalizable, as they may be enriched for health-councious internet-connected participants, and thus underestimate disparities in at-risk demographic groups. We show that symptom-tracking app participants and those with smartphones have higher testing rates than all UMD-Facebook survey respondents.

Confounding and measurement bias in this observational study using self-reported covariates and outcomes may also cause us to miss other important issues related to testing. We adjusted for common confounders, and attempted to identify proxies for the knowledge gap rather than attribute causality. There is no timely, efficient trial to conduct analyses of this scale. Self-report could introduce non-differential and differential measurement error, including the possibility of some events being omitted, or recorded inaccurately or inappropriately. Furthermore, the financial implications of having to self-isolate disproportionately affect the poorest, and may increase unwillingness to test^33^ and respondents may be wary of self-reporting socially stigmatized reasons for not complying with guidance. Lastly, selection can theoretically induce collider bias^34^ if the exposure and outcome are both causes of participation or subpopulation selection.

## Conclusion

Testing is a fundamental principle of population-wide transmission mitigation. While the UK now has sufficient testing capacity, consistent guidelines, and free testing for those who qualify that is coordinated centrally, still we see a 25% testing gap among those with test-qualifying symptoms. We show this gap may be driven in part by a lack of understanding of mild COVID-19, national testing criteria, and testing access, especially among the elderly and those who have had fewer years of education. We propose altering the course of the UK testing programme to address this knowledge barrier to COVID-19 testing. In addition, other countries may benefit from improved understanding of modifiable barriers.

## Data Availability

Zoe Platform data used in this study is available to researchers through UK Health Data Research using the following link:
https://web.www.healthdatagateway.org/dataset/fddcb382-3051-4394-8436-b92295f14259
Requests for UMD/Facebook data may be completed through: https://dataforgood.fb.com/docs/covid-19-symptom-survey-request-for-data-access/

## Authors and Contributions

MSG, CMA, TV, and CJS conceived of the study. CMA and MSG wrote the first draft of the manuscript. MSG, AM, CH, JC oversaw acquisition of Zoe data collection. MSG, TV, CMA, ATC, TDS, SO, CJS contributed to the Zoe follow-up survey design. CMA and JSB verified the underlying UMD-Facebook data. MSG and TV verified the underlying KCL/Zoe data. CMA and MSG contributed to the analyses. All authors contributed to the interpretation of the data for the work, revised the manuscript, and approved the final manuscript.

## Acknowledgments

CMA and JSB: Facebook Sponsored Research Agreement [INB1116217]. CHS Alzheimer’s Society Junior Fellowship [AS-JF-17-011]. Support for the COVID Symptom Study (UK data) was provided by the NIHR-funded Biomedical Research Centre based at GSTT NHS Foundation Trust. This work was supported by the UK Research and Innovation London Medical Imaging & Artificial Intelligence Centre for Value Based Healthcare. ZOE Global provided in kind support for all aspects of building, running and supporting the app and service to all users worldwide. Support for this study was provided by the NIHR-funded Biomedical Research Centre based at GSTT NHS Foundation Trust. Investigators also received support from the Wellcome Trust (212904/Z/18/Z, WT203148/Z/16/Z), the MRC/BHF (MR/M016560/1), Alzheimer’s Society, EU, NIHR, CDRF, and the NIHR-funded BioResource, Clinical Research Facility and BRC based at GSTT NHS Foundation Trust in partnership with KCL, the UK Research and Innovation London Medical Imaging & Artificial Intelligence Centre for Value Based Healthcare, the Wellcome Flagship Programme (WT213038/Z/18/Z), the Chronic Disease Research Foundation, and DHSC. ATC was supported in this work through a Stuart and Suzanne Steele MGH Research Scholar Award. The Massachusetts Consortium on Pathogen Readiness (MassCPR) and Mark and Lisa Schwartz supported MGH investigators (DAD, LHN, ATC).

## Declaration of Interests

The authors CMA, BR and JSB declare no competing interests. AM, JCP, CH, JW are employees of Zoe Global Ltd. TDS is a consultant to Zoe Global Ltd. DAD and ATC previously served as investigators on a clinical trial of diet and lifestyle using a separate smartphone application that was supported by Zoe Global. ATC reports grants from Massachusetts Consortium on Pathogen Readiness, during the conduct of the study; personal fees from Pfizer Inc., personal fees from Boehringer Ingelheim, personal fees from Bayer Pharma AG, outside the submitted work. DAD reports grants from National Institutes of Health, grants from MassCPR, grants from American Gastroenterological Association during the conduct of the study.

## Data Availability

Zoe Platform data used in this study is available to researchers through UK Health Data Research using the following link: https://web.www.healthdatagateway.org/dataset/fddcb382-3051-4394-8436-b92295f14259

Requests for UMD/Facebook data may be completed through: https://dataforgood.fb.com/docs/covid-19-symptom-survey-request-for-data-access/

## Role of the funding source

The funding sources played no role in the study design, collection, analysis, interpretation, writing or decision to submit the paper for publication.

## Ethics & IRB

Boston Children’s Hospital IRB (P00023700) to use UMD-Facebook data. King’s College London ethics committee (REMAS ID 18210; LRS-19/20-18210) to use Zoe data.

## Supplementary Material

### S1 Supplementary Table: Zoe CSS questions

In each daily report, users are asked about any symptoms they experience that day, and any COVID-19 tests they have had.

### Symptoms

Users are asked “How do you feel physically right now?”. If they select “I’m not feeling quite right”, they are presented with a symptom options checklist. The symptom options specifically analysed in this study are:

Are you experiencing any of the below symptoms?

□ Fever or feel too hot
□ Loss of smell / taste
□ Altered smell / taste (things smell or taste different to usual)
□ Persistent cough (coughing a lot for more than an hour, or 3 or more coughing episodes in 24 hours)

### Testing

Users are shown a list of all COVID-19 tests they have logged through the app. They are able to add new tests, edit existing entries, or select “This list is correct”. If a user chooses to add a new test, they are asked:

- Do you know the date of your test?

- If yes, select date
- How was the test performed?

- A swab of my nose or throat
- I spat in a cup/tube
- A finger-prick blood test
- A blood test, done using a needle
- Other, please specify (free text)
- Where was this test performed?

- At Home
- Drive-through Regional Testing Centre
- Hospital (not drive-through)
- GP
- Chemist / Pharmacy
- Work (excluding hospital or GP)
- Other, please specify
- What are the results of this test?

- Negative
- Positive
- Not clear/ failed
- Waiting for results

### S2 Supplementary Table: Zoe CSS testing survey questions

#### Q1 Have you experienced any of the following symptoms in the last month? (check all that apply)

□ Fever
□ Persistent cough
□ Loss of smell or taste
□ Altered smell or taste
□ Shortness of breath
□ Fatigue
□ Muscle or body aches
□ Headache
□ Sore throat
□ Congestion or runny nose
□ Nausea or vomiting
□ Diarrhea
□ None of the above

If None of the above; Proceed to **Q2a** then **SURVEY END**.

Else; Proceed to **Q2.**

#### Q2 Did these symptoms qualify you for a COVID-19 swab test where you live?

□ Yes
□ No
□ Do not know

If YES; Proceed to **Q3**

If NO or DO NOT KNOW; Proceed to **Q2a**

#### SQ2a What symptoms qualify you for a covid test where you live? (check all that apply)

□ Fever
□ Persistent cough
□ Loss of smell or taste
□ Altered smell or taste
□ Shortness of breath
□ Fatigue
□ Muscle or body aches
□ Headache
□ Sore throat
□ Congestion or runny nose
□ Nausea or vomiting
□ Diarrhea
□ None of the above
□ Do not know

Proceed to **Q3** (if arrived here directly from Q1, proceed to **SURVEY END**)

#### Q3 Did you try to get a swab test?

If YES; Proceed to **Q4**

If NO; Proceed to **Q3A**

##### Q3a Please select the reason(s) you did not attempt to get a swab test (all that apply)

□ I did not believe I could/should get a test with my symptoms at the time
□ My symptoms were normal / not new for me
□ I thought that travelling to a testing appointment would be difficult or risky
□ I was concerned about discomfort or pain from the swab
□ I have already had Covid and did not think I could get it again
□ I couldn’t find the time to go to an appointment (e.g. couldn’t get time off work/childcare)
□ I was concerned testing positive might affect me financially (job/income/studies)
□ I was concerned about the cost of the the test
□ I was concerned testing positive would affect me socially (time away from friends/family)
□ I was concerned about testing positive and being contacted by contact tracers or health authorities
□ The tests are not reliable
□ OTHER - free text

Proceed to **SURVEY END**

#### Q4 Did you receive a COVID-19 test (swab or otherwise) within 14 days of having such symptoms?

If YES; Proceed to **Q5**

If NO, Proceed to **Q4a**

##### Q4a TRIED TO GET TEST REASONS

Please state the reason(s) the test did not happen (all that apply): I did not know how to get a test

□ There were no testing appointments available
□ There were no home testing kits available
□ The test never arrived/I never received the result back
□ I thought that travelling to a testing appointment would be too risky
□ I was concerned about swabbing myself/being swabbed
□ Difficult with transportation to my appointment (e.g. too far away/I didn’t have a vehicle)
□ I couldn’t find the time to go to an appointment (e.g. couldn’t get time off work/childcare)
□ I could not afford the test
□ I was told by a doctor or testing centre that I didn’t need a test
□ OTHER - free text

Proceed to **SURVEY END**

### Q5 What date was your COVID-19 test? Please also log this test through the app, if you have not done so already

[Date entry field]

Proceed to **SURVEY END SURVEY END**

---

**S3 Supplementary Table:**
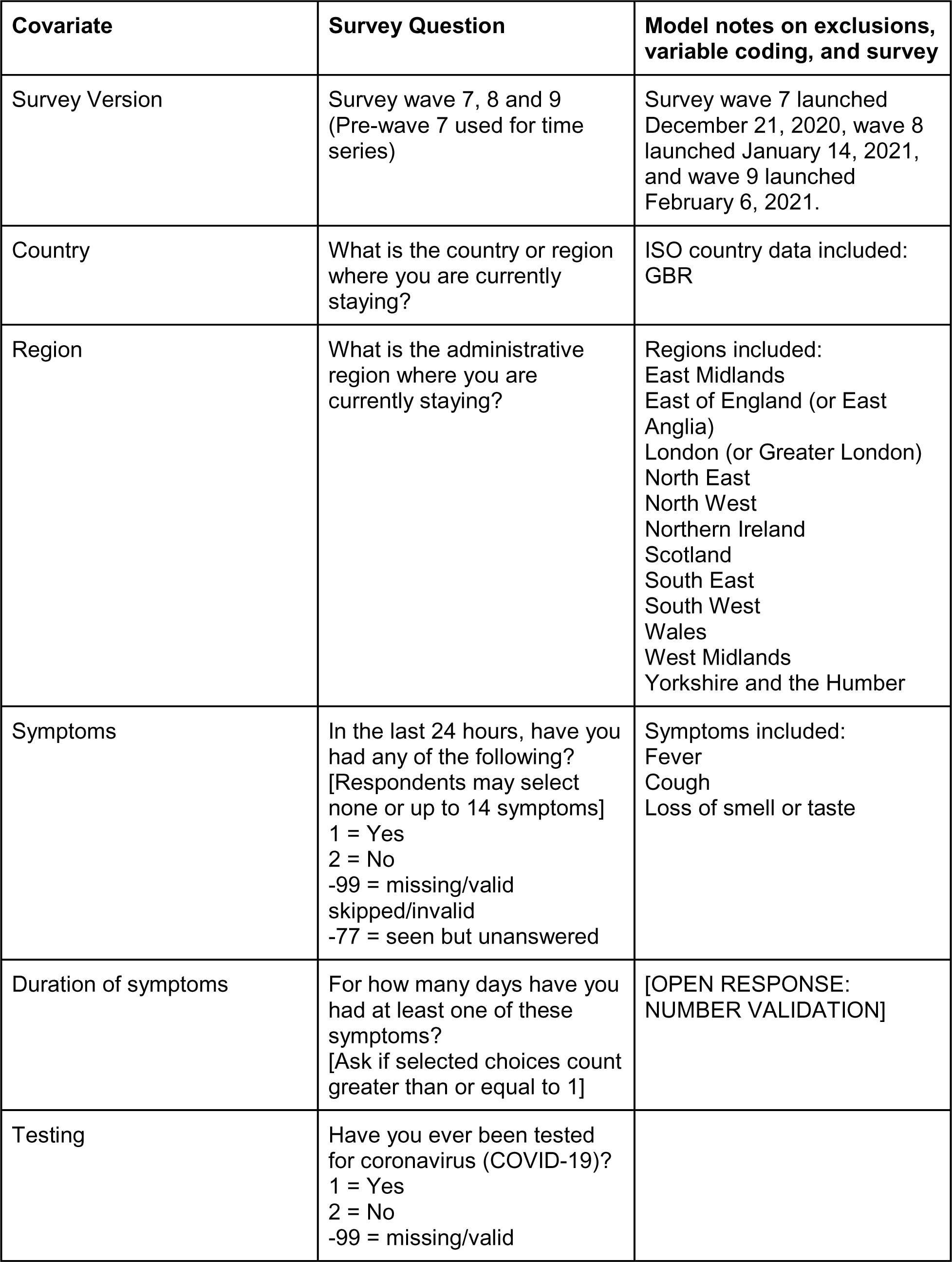
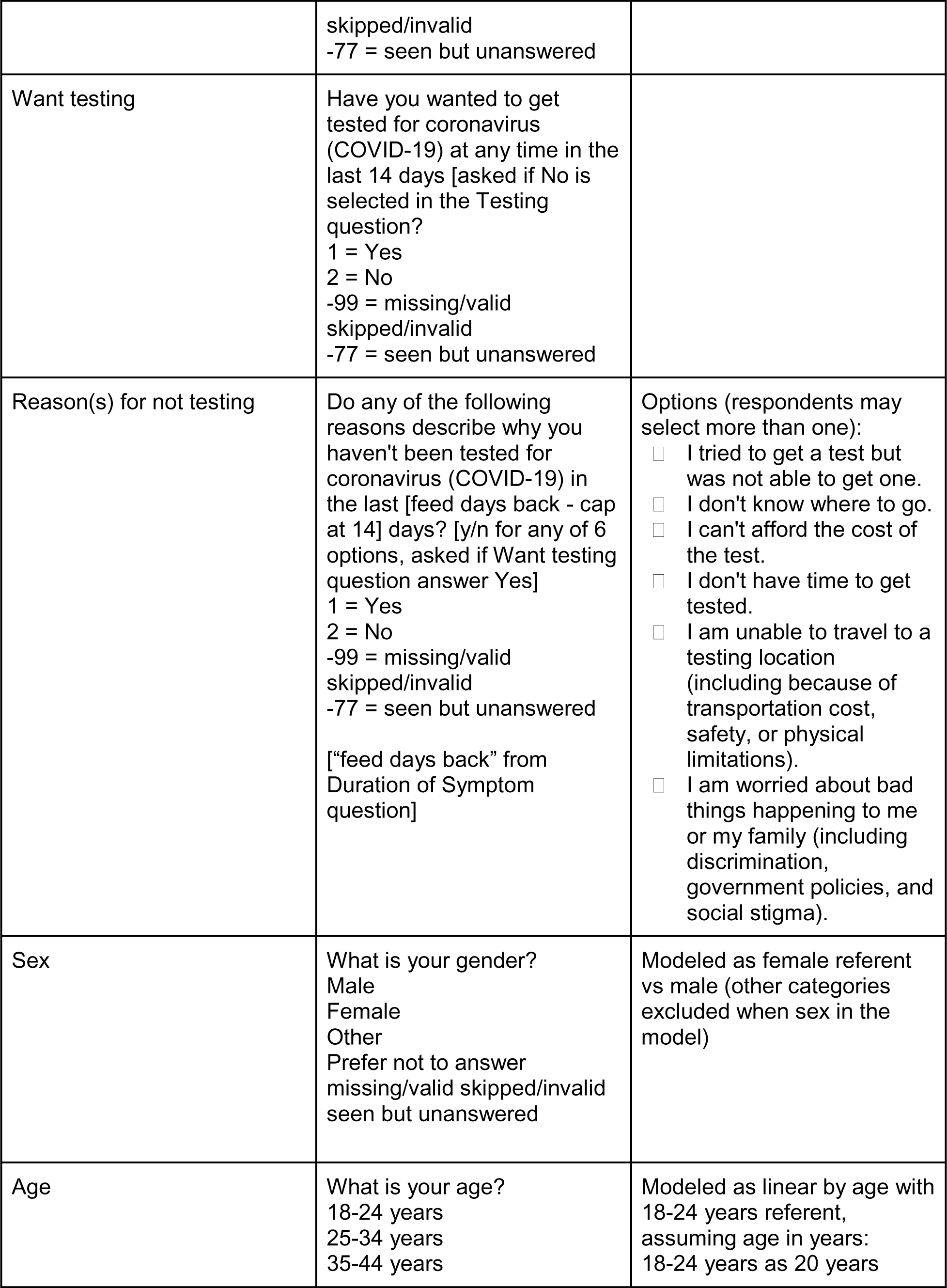
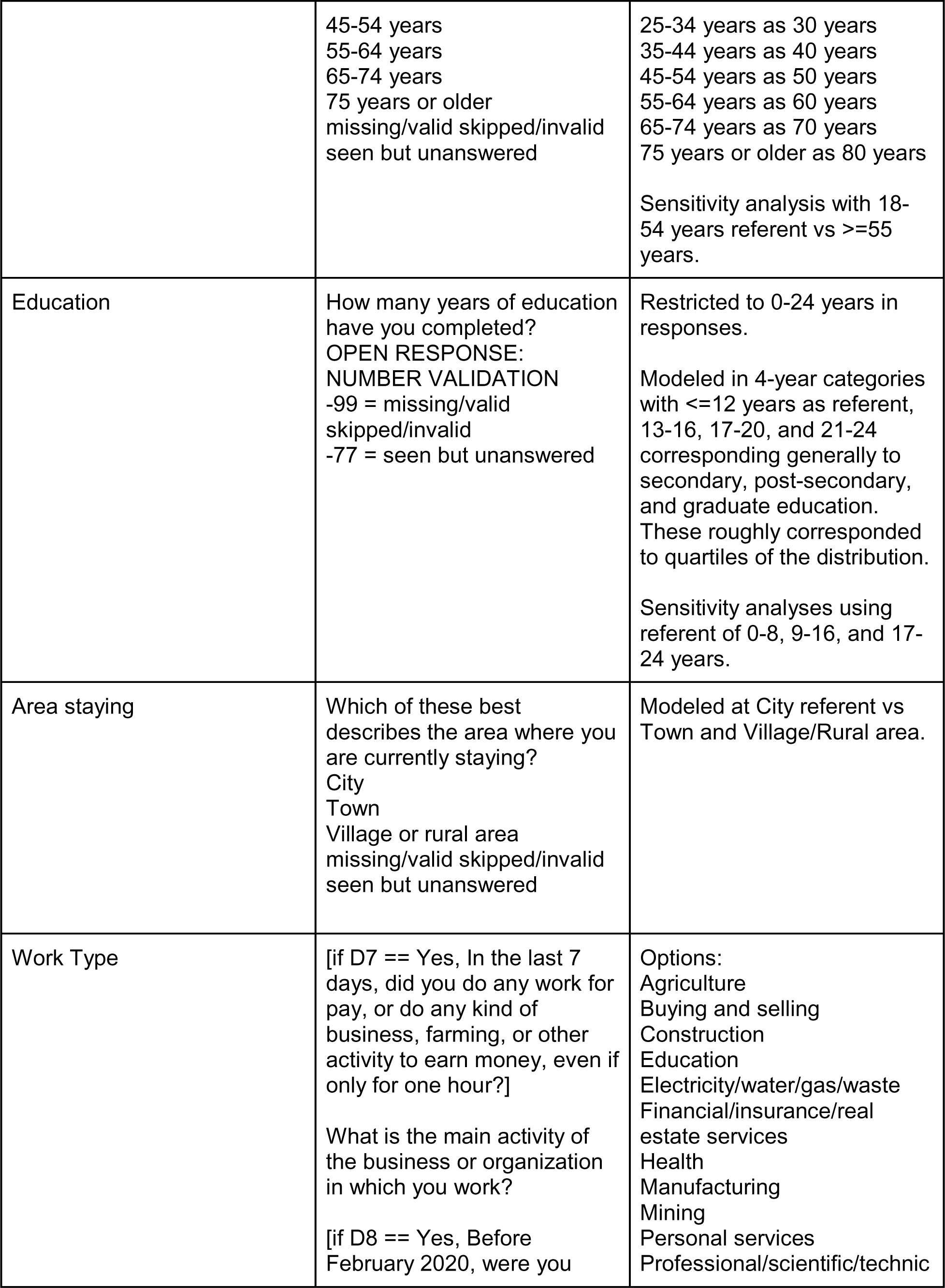
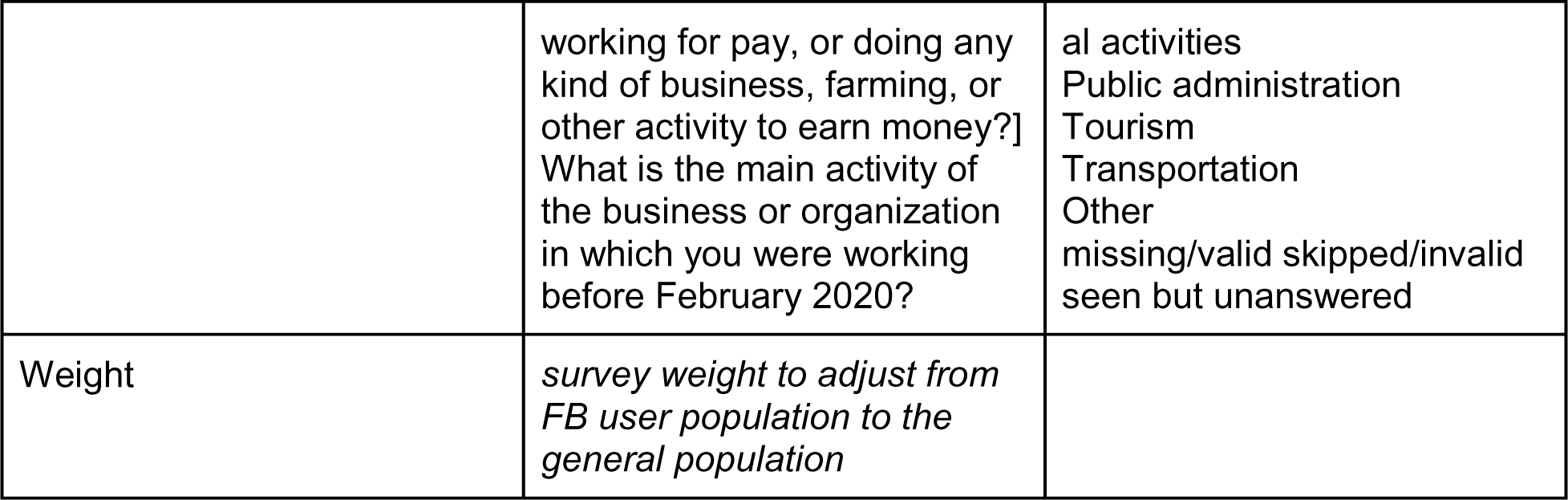
UMD Facebook Survey questions.

**S4 Supplementary Table:**
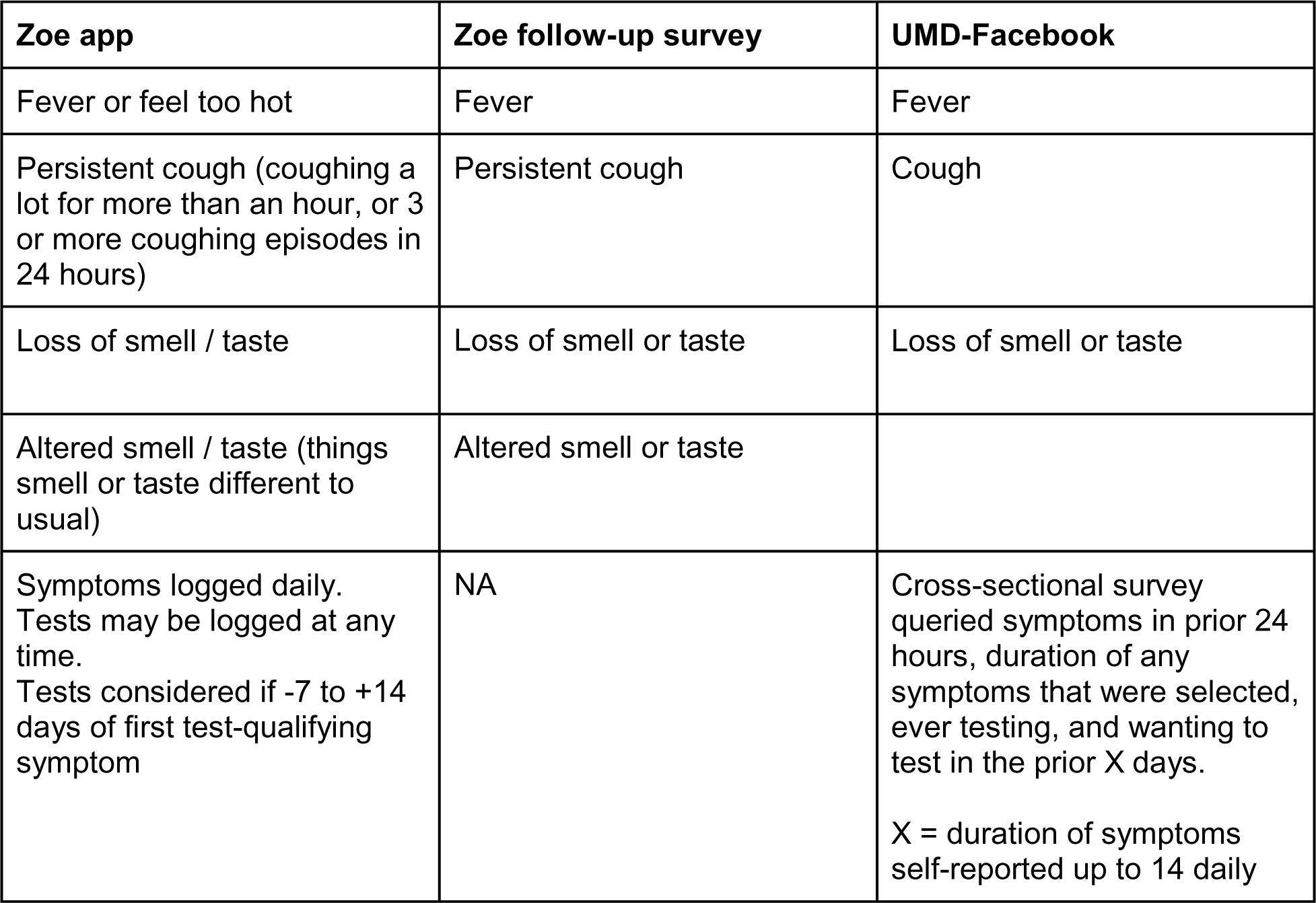
Comparison of question wording between Zoe and UMD-Facebook. Comparison for all symptoms considered as test-qualifying in this work.

**S6 Supplementary Table:**
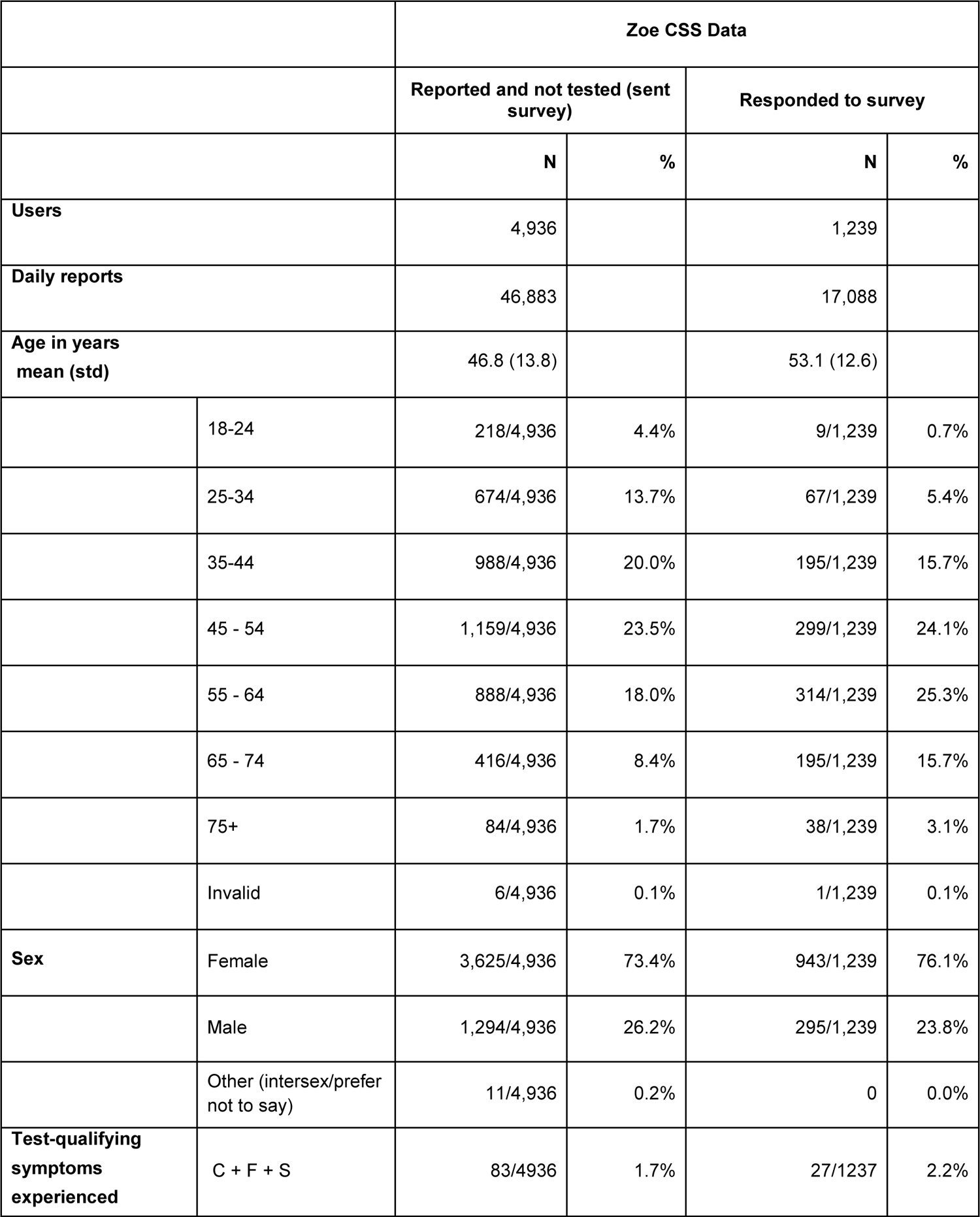
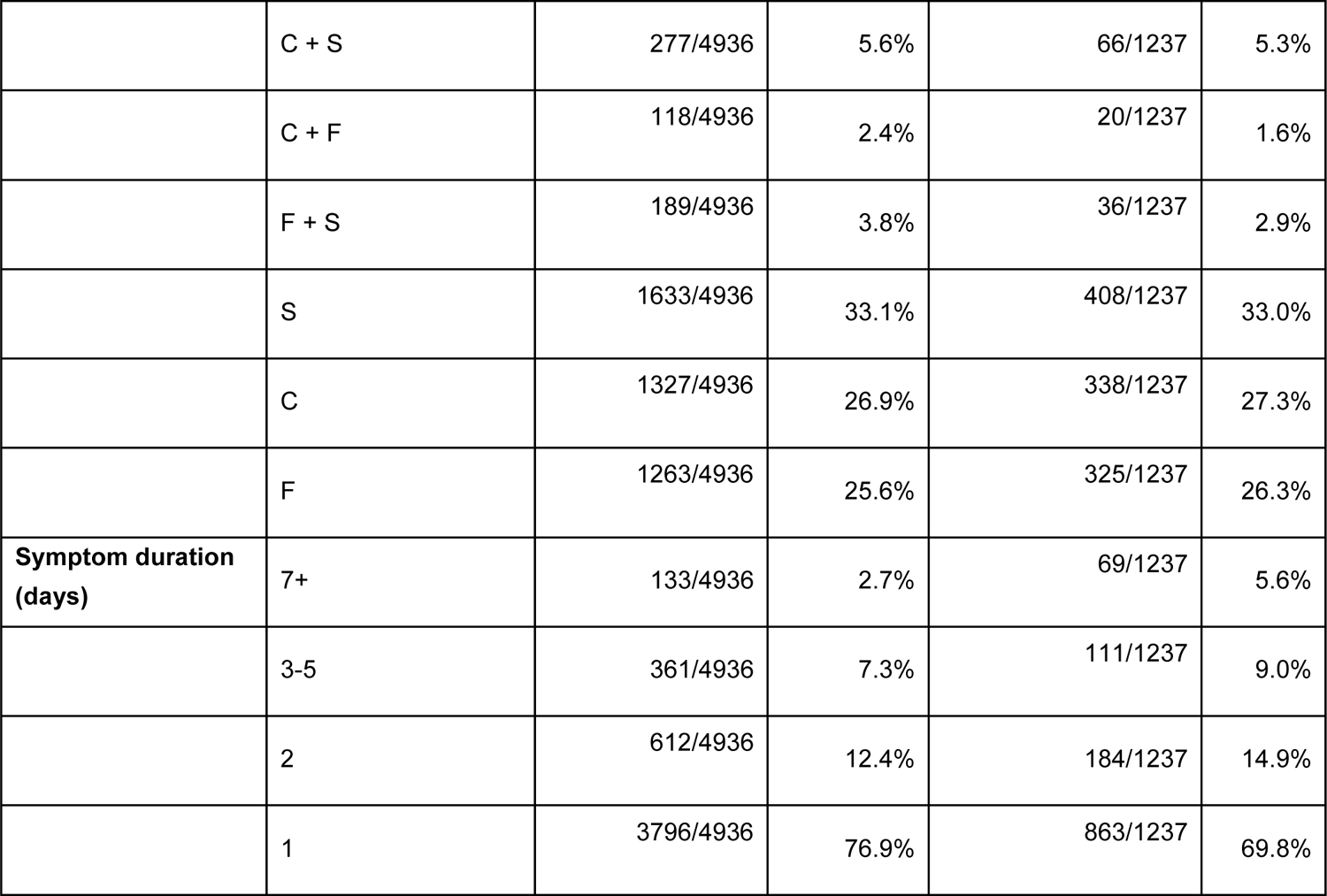
Comparison of Zoe survey respondents to all surveyed. Comparison between all users sent the Zoe follow-up survey and respondents.

**S7 Supplementary Table:** Ability to recall symptoms in Zoe CSS survey. Logistic regression models for the association of ability to recall test-qualifying symptoms, including covariates of sex, age, symptom duration, number of test-qualifying symptoms, and time between symptom onset and survey response.

|  |  | Odds Ratios |  |  |  |  |
| --- | --- | --- | --- | --- | --- | --- |
| Model | Number | Sex | Age | Symptom duration | Number test-qualifying symptoms | Time from symptom onset |
| Univariate Sex (Female Referent) | 1 | 1.048 (0.980-1.121) p=0.170 | --- | --- | --- | -- |
| Univariate Age (Per Decade Above 18-24 Referent) | 2 | --- | 1.005 (0.982-1.028) p=0.660 | --- | --- | --- |
| Univariate Symptom duration (Per day above 0) | 3 | --- | --- | **1.065 (1.054-1.076) p<0.001** | --- | --- |
| Univariate number of test-qualifying symptoms (1 Referent) | 4 | --- | --- | --- | **1.302 (1.220-1.391) p<0.001** | --- |
| Univariate time from symptom onset to survey (Per day, 10 days referent) | 5 | --- | --- | --- | --- | **0.995 (0.991-1.000) p=0.029** |
| Multivariate | 6 | 0.947 (0.888-1.009) p=0.091 | 0.997 (0.975-1.018) p=0.760 | **1.062 (1.049-1.074) p<0.001** | **0.991 (0.987-0.994) p<0.001** | **1.142 (1.067-1.223) p=0.000** |

**S8 Supplementary Table:** Understanding of testing criteria. Lgistic regression models for the association of ability to recall test-qualifying symptoms, including covariates of sex, age, index of multiple deprivation, rural-urban classification

| Symptoms | Model | Number | Odds Ratios |  |  |  |
| --- | --- | --- | --- | --- | --- | --- |
|  |  |  | Sex | Age | IMD | RUC |
| All three (fever, cough, loss of smell) | Univariate Sex (Female Referent) | 1 | 0.927 (0.853-1.007) p=0.072 | --- | --- | --- |
|  | Univariate Age (Per Decade Above 18-24 Referent) | 2 | --- | **0.908 (0.883-0.933) p=0.000** | --- | --- |
|  | Univariate IMD (By tercile, referent = 1, most deprived) | 3 | --- | --- | 1.002 (0.953-1.053) p=0.944 | --- |
|  | Univariate rural/urban classification (Rural referent) | 4 | --- | --- | --- | 0.976 (0.894-1.066) p=0.587 |
|  | Multivariate | 5 | 0.969 (0.894-1.052) p=0.455 | **0.906 (0.881-0.932) p<0.001** | 1.018 (0.970-1.069) p=0.465 | 0.943 (0.865-1.028) p=0.181 |
| Fever | Univariate Sex (Female Referent) | 1 | **0.915 (0.844-0.993) p=0.033** | --- | --- | --- |
|  | Univariate Age (Per Decade Above 18-24 Referent) | 2 | --- | **0.911 (0.887-0.936) p<0.001** | --- | --- |
|  | Univariate IMD (By tercile, referent = 1, most deprived) | 3 | --- | --- | 1.007 (0.959-1.057) p=0.785 | --- |
|  | Univariate rural/urban classification (Rural referent) | 4 | --- | --- | --- | 0.965 (0.885-1.052) p=0.419 |
|  | Multivariate | 5 | 0.955 (0.882-1.035) p=0.264 | **0.910 (0.885-0.935) p<0.001** | 1.022 (0.975-1.072) p=0.365 | 0.934 (0.859-1.017) p=0.114 |
| Persistent cough | Univariate Sex (Female Referent) | 1 | 0.937 (0.865-1.014) p=0.105 | --- | --- | --- |
|  | Univariate Age (Per Decade Above 18-24 Referent) | 2 | --- | **0.909 (0.885-0.933) p<0.001** | --- | --- |
|  | Univariate<br>IMD(By tercile,<br>referent = 1,<br>most deprived) | 3 | --- | --- | 0.984 (0.938-1.032) p=0.506 | --- |
|  | Univariate<br>rural/urban<br>classification<br>(Rural referent) | 4 | --- | --- | --- | 1.022 (0.939-1.111) p=0.615 |
|  | Multivariate | 5 | 0.980 (0.906-1.059) p=0.604 | **0.909 (0.885-**<br>**0.934) p<0.001** | 1.002 (0.956-1.049) p=0.94 | 0.986 (0.908-1.071) p=0.742 |
| Loss of<br>taste/smell | Univariate Sex<br>(Female<br>Referent) | 1 |  | --- | --- | --- |
|  | Univariate Age<br>(Per Decade<br>Above 18-24<br>Referent) | 2 | --- | **0.910 (0.885-**<br>**0.934) p<0.001** | --- | --- |
|  | Univariate<br>IMD(By tercile,<br>referent = 1,<br>most deprived) | 3 | --- | --- | 1.002 (0.954-1.051) p=0.948 | --- |
|  | Univariate<br>rural/urban<br>classification<br>(Rural referent) | 4 | --- | --- | --- | 1.017 (0.934-1.108) p=0.698 |
|  | Multivariate | 5 | 0.973 (0.899-1.053) p=0.501 | **0.909 (0.885-**<br>**0.935) p<0.001** | 1.020 (0.972-1.069) p=0.422 | 0.984 (0.905-1.070) p=0.710 |
| Altered<br>taste/smell | Univariate Sex<br>(Female<br>Referent) | 1 | 1.045 (0.968-1.129) p=0.258 | --- | --- | --- |
|  | Univariate Age<br>(Per Decade<br>Above 18-24<br>Referent) | 2 | --- | 0.978 (0.952-1.004) p=0.091 | --- | --- |
|  | Univariate<br>IMD(By tercile,<br>referent = 1,<br>most deprived) | 3 | --- | --- | 0.985 (0.940-1.031) p=0.513 | --- |
|  | Univariate<br>rural/urban<br>classification<br>(Rural referent) | 4 | --- | --- | --- | 1.027 (0.947-1.114) p=0.524 |
|  | Multivariate | 5 | 1.058 (0.979-1.144) p=0.153 | 0.976 (0.950-1.003) p=0.075 | 0.990 (0.945-1.037) p=0.657 | 1.017 (0.937-1.104) p=0.686 |

**S9 Supplementary Table:** Knowing where to test. Logistic regression models for the association of not knowing where to test, including covariates of age, sex, education and where living, comparing those who answered “yes” and “no”. Further sensitivity analyses assuming missing responses to be “no” lowers the proportion of answered “yes” and attenuates p-values, while effect estimates are directionally consistent and statistically significant (e.g. education multivariate model per 4 years of education, OR=0.711 (0.619-0.813), p=8.84e-07). Modeling age as young (referent 18-54 years) vs old (55+), produced a similar estimate of the association of age (OR=2.1 (1.65, 2.69), p=2.12e-09) with not knowing where to test. Modeling education in 8-year categories starting prior to completion of secondary school (0-8, 9-16, and 17-24) was also similar OR=0.662, 0.528-0.830, p=3.65e-4). See Supplementary Figure 7 and 9 for additional sensitivity analyses and qualitative investigations.

| Model | Number | (Intercept) | Sex<br>(Female<br>Referent) | Age in<br>Decade<br>(18-24 Years<br>Referent) | Education in<br>4 Years<br>(≤12 Years<br>Referent) | Where Living<br>(City<br>Referent) |
| --- | --- | --- | --- | --- | --- | --- |
| Univariate<br>Sex | 1 | OR=0.65<br>(0.554-0.762)<br>$p=1.228e-07$ | OR=1.334<br>(1.064-1.675)<br>$p=1.277e-02$ | NA | NA | NA |
| Univariate<br>Age | 2 | OR=0.327<br>(0.236-0.45)<br>$p=1.220e-11$ | NA | OR=1.207<br>(1.129-1.292)<br>$p=5.007e-08$ | NA | NA |
| Univariate<br>Education | 3 | OR=1.069<br>(0.915-1.25)<br>$p=3.997e-01$ | NA | NA | OR=0.648<br>(0.56-0.747)<br>$p=3.281e-09$ | NA |
| Univariate<br>Where Living | 4 | OR=0.653<br>(0.519-0.817)<br>$p=2.186e-04$ | NA | NA | NA | OR=1.201<br>(0.926-1.562)<br>$p=1.696e-01$ |
| Multivariate<br>Education<br>Adjusting for<br>Age and Sex | 5 | OR=0.417<br>(0.288-0.601)<br>$p=3.101e-06$ | OR=1.218<br>(0.962-1.543)<br>$p=1.009e-01$ | OR=1.203<br>(1.119-1.294)<br>$p=5.456e-07$ | OR=0.655<br>(0.563-0.759)<br>$p=3.051e-08$ | NA |
| Education<br>with Survey<br>Weights | 6 | OR=1.099<br>(0.904-1.336)<br>$p=3.433e-01$ | NA | NA | OR=0.672<br>(0.564-0.799)<br>$p=7.842e-06$ | NA |

**S5 Supplementary Figure:**
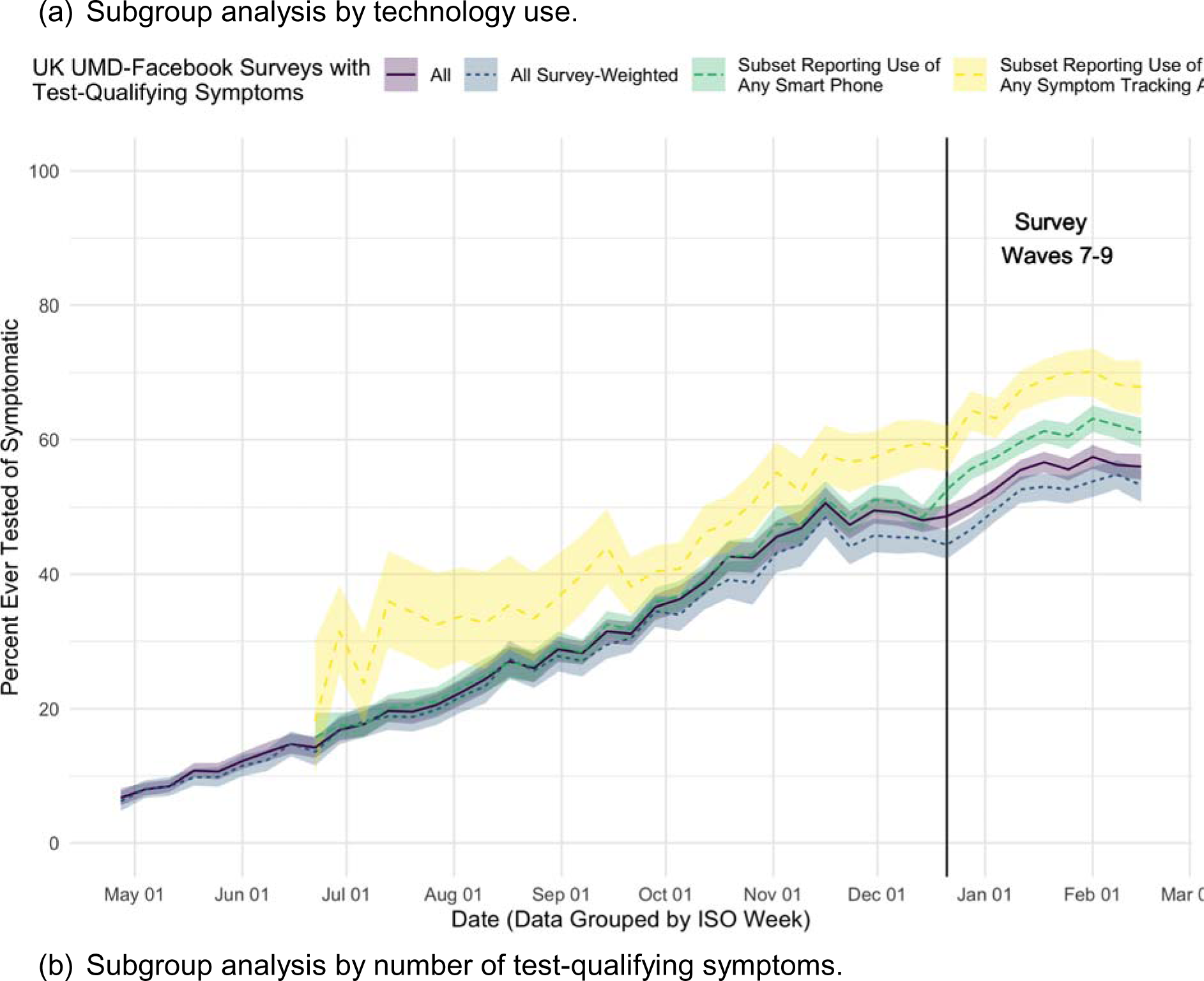
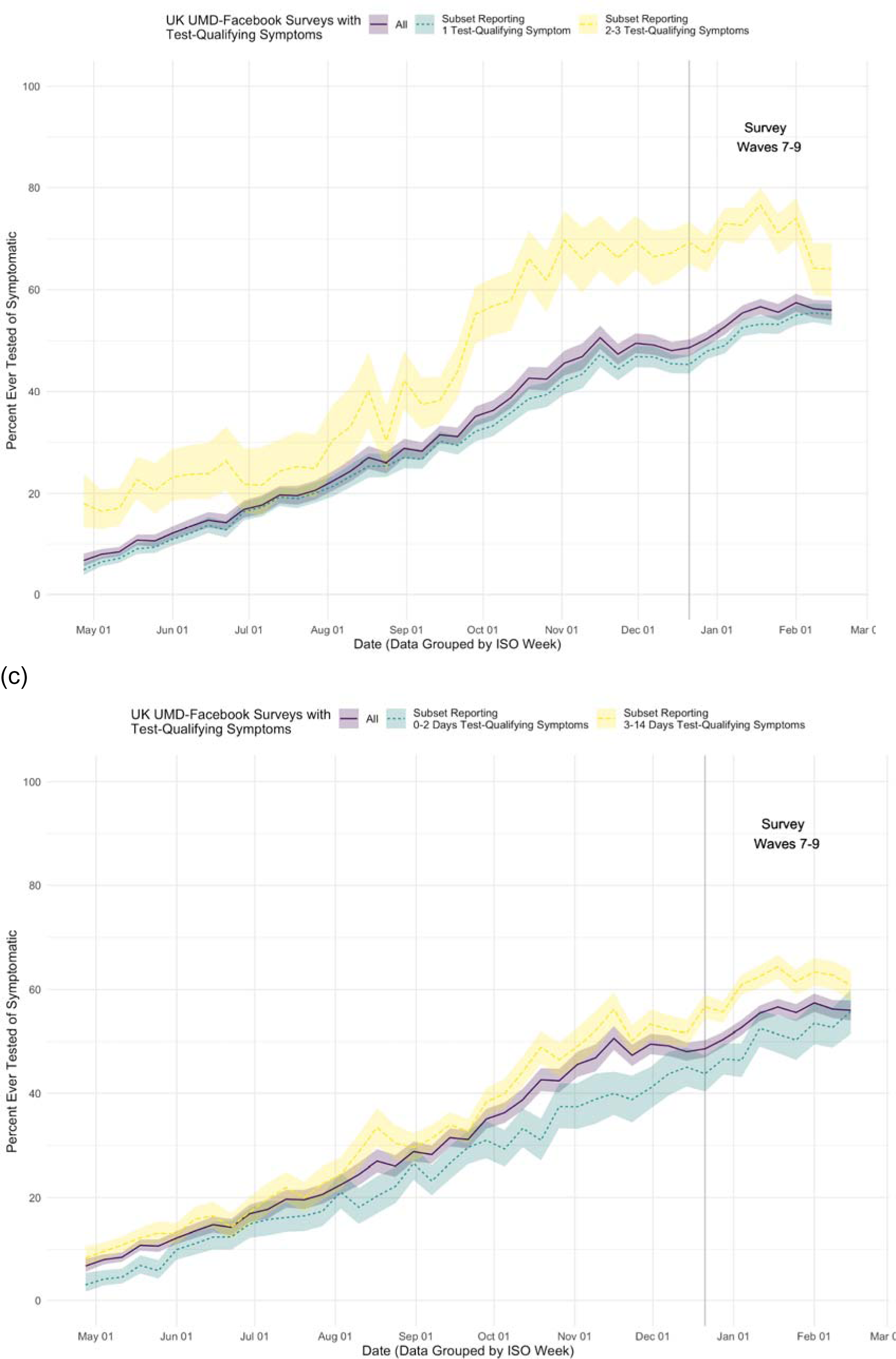
Testing trends. UMD Facebook UK COVID-19 Symptom Survey temporal trends in the proportion of symptomatic survey respondents ever tested from survey start (April 30, 2020). Additional questions for the never tested respondents were added in survey wave 7 and beyond (December 21, 2020, vertical grey line). Surveys with non-missing geographic region in the UK, self-reporting at least one of fever, cough or loss of smell/taste in the prior 24 hours (N=107,021, solid purple), plotting proportion (Wilson method for binomial 95% confidence intervals) who indicate ever testing for COVID-19. (a) Starting June 27, 2020, respondents were queried about technology use. There were N=67508 (63.1%, dashed green) who reported using a smartphone device, while N=16,488 (15.4%, dashed yellow) reported using any symptom tracking app. Survey-weighted mean (+/- 2 standard deviation) for all surveys with test-qualifying symptoms included for comparison. This was generally lower than the raw proportion ever tested. Testing proportion among cross-sectional surveys varied by (b) number of test-qualifying symptoms and (c) duration of symptoms (for those who reported symptom duration), similar to the prospective, longitudinal Zoe app findings.

**S10 Supplementary Figure:**
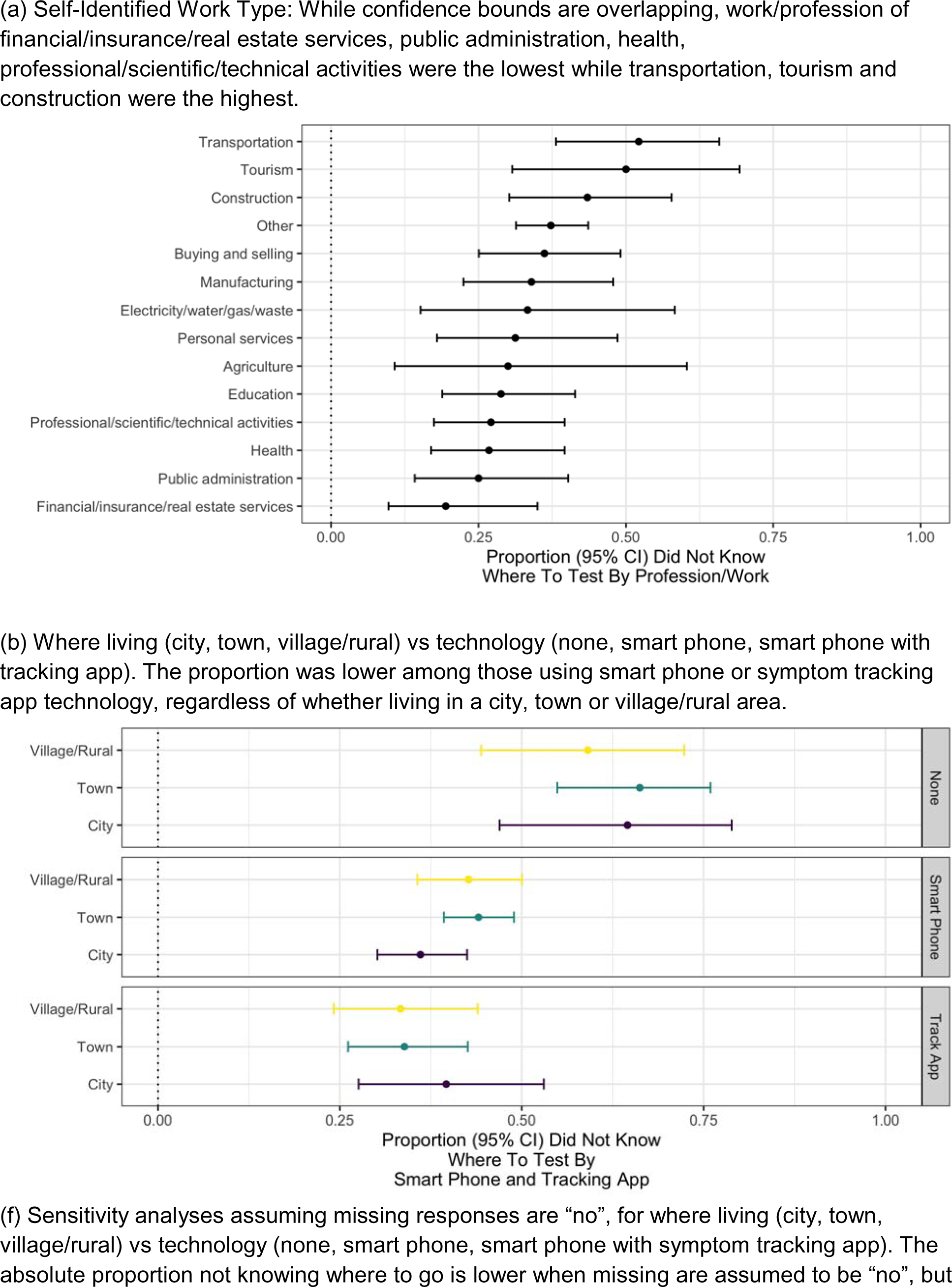
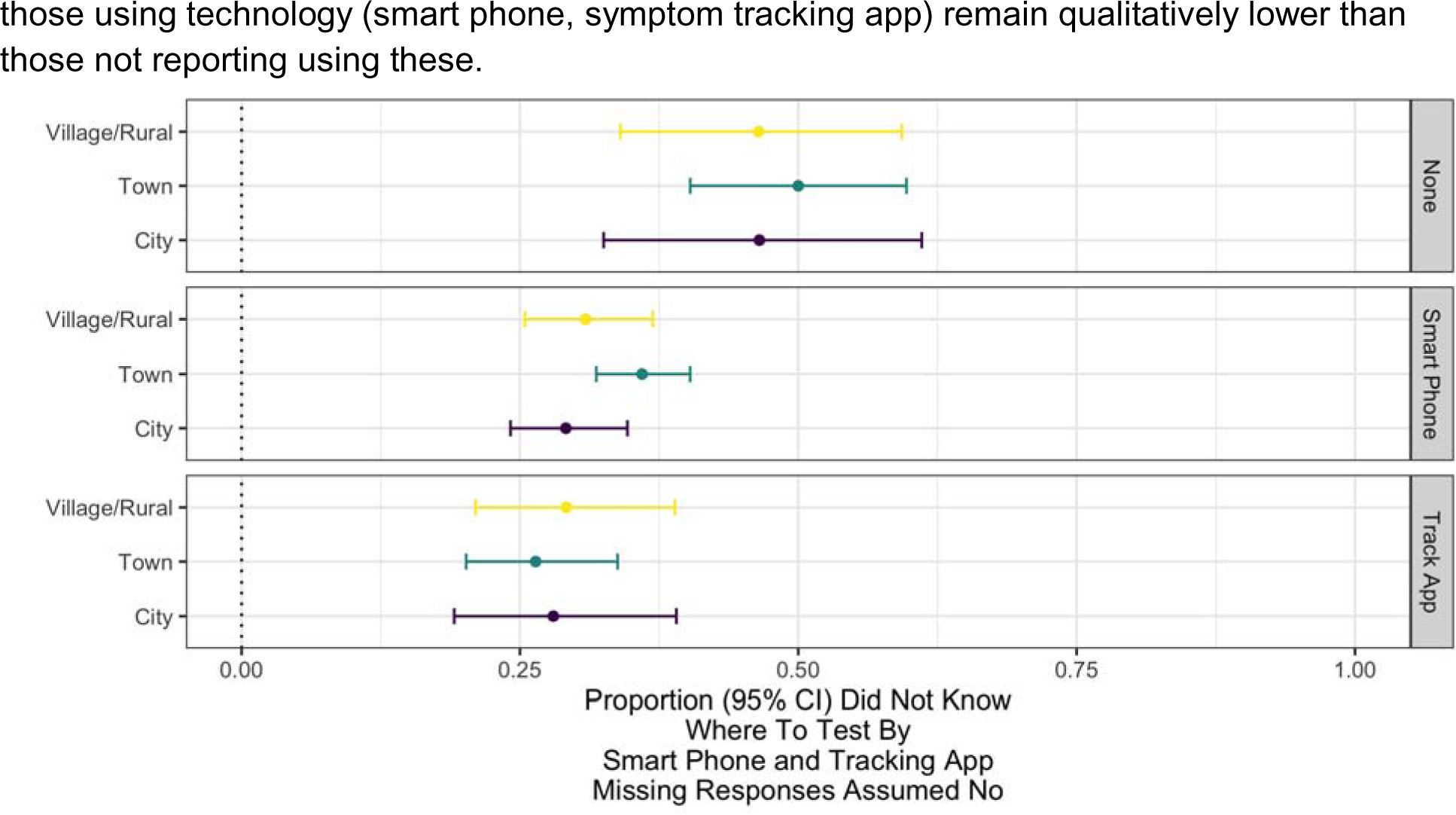
Qualitative demographic-knowledge relationships. Shown are the proportion (Wilson 95% confidence interval for binomial) reporting they did not know where to test (“yes” vs “yes” and “no”) across various demographic factors including: a) self-identified work type, b) where living (city,town, village/rural) vs technology used (none, smart phone, symptom tracking app). Missing responses are excluded in the primary analyses. c) Sensitivity analysis assuming missing responses are “no”, and are generally lower but with the same demographic patterns.

**S11 Supplementary Figure:**
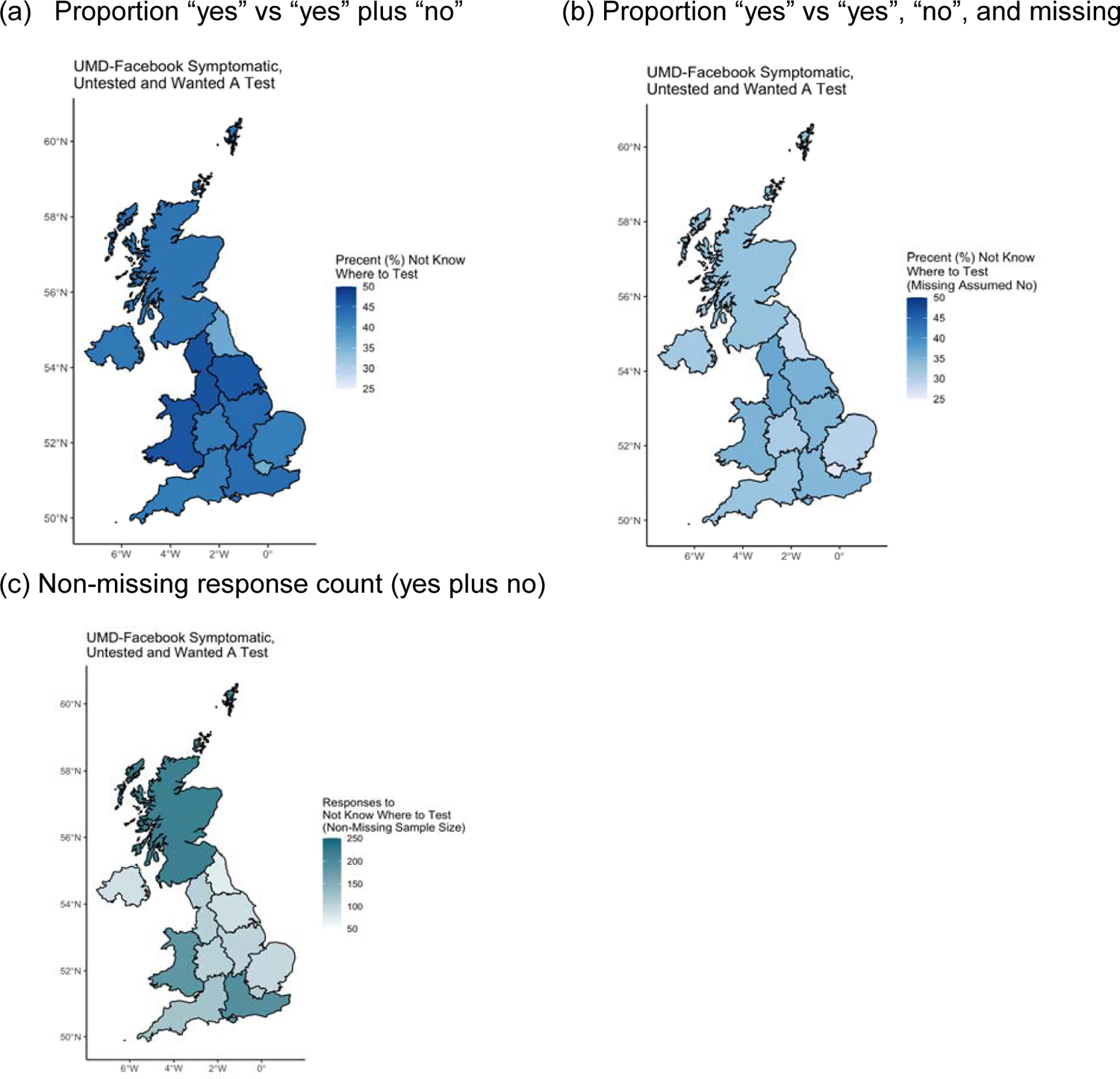
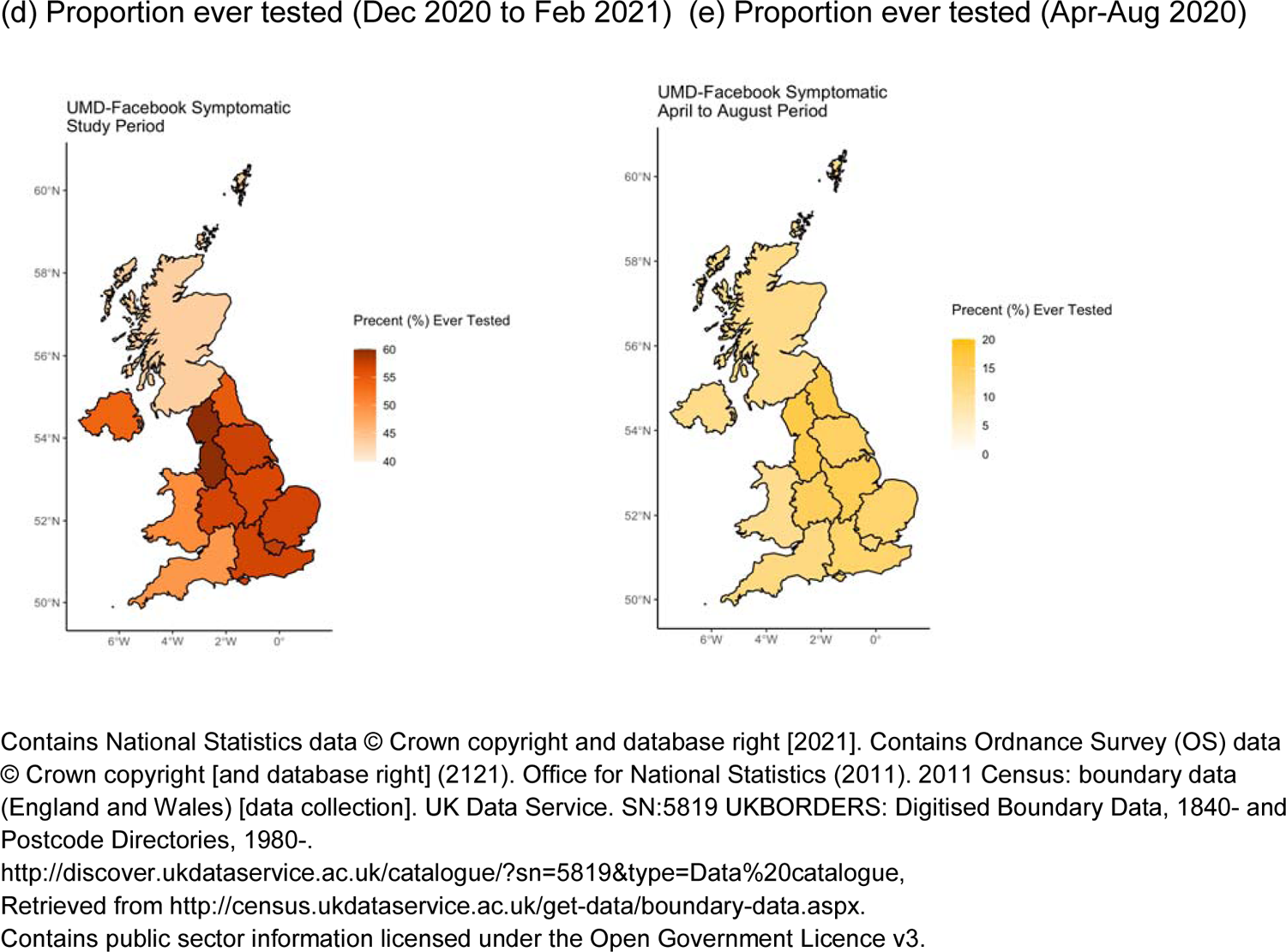
Participant locations for UMD-Facebook Survey. Map of proportion of symptomatic, untested survey respondents who wanted to test but did not know where to test (a) proportion, (b) sensitivity analysis assuming missing responses were “no”, and (c) number of surveys with yes or no responses to the reason “I don’t know where to go” when asked “Do any of the following reasons describe why you haven’t been tested for coronavirus (COVID-19) in the last X days? [y/n]”. X is the self-reported duration of symptoms (up to 14 days) and (d). (d) Geographic distribution of proportion of symptomatic ever tested during the study period and (e) early in the pandemic (Apr-Aug 2020). Not the orange (d) and yellow (e) color scales span the same percentage but have different minimum values. Early in the pandemic the testing gap was very similar across geographic areas. During the study period, the testing gap is closing in all geographic areas, with qualitatively better testing gains in the testing rates in some areas.

## Notes

### Summary of Updates

Fixed formatting issues.

